# Characterization of SARS-CoV-2 intrahost genetic evolution in vaccinated and non-vaccinated patients from the Kenyan population

**DOI:** 10.1101/2025.03.03.25323296

**Authors:** Doreen Lugano, Kennedy Mwangi, Bernard Mware, Gilbert Kibet, Shebbar Osiany, Edward Kiritu, Paul Dobi, Collins Muli, Regina Njeru, Tulio de Oliveira, M. Kariuki Njenga, Andrew Routh, Samuel O. Oyola

## Abstract

Vaccination is a key control measure of COVID-19 by preventing severe effects of disease outcomes, reducing hospitalization rates and death, and increasing immunity. However, vaccination can affect the evolution and adaptation of SARS-CoV-2, largely through vaccine-induced immune pressure. Here we investigated intrahost recombination and single nucleotide variations (iSNVs) on the SARS-CoV-2 genome in non-vaccinated and vaccinated sequences from the Kenyan population to profile intrahost viral genetic evolution and adaptations driven by vaccine-induced immune pressure. We identified recombination hotspots in the S, N, and ORF1a/b genes and showed the genetic evolution landscape of SARS-CoV-2 by comparing within-wave and inter-wave recombination events from the beginning of the pandemic (June 2020) to (December 2022) in Kenya. We further reveal differential expression of recombinant RNA species between vaccinated and non-vaccinated individuals and perform an in-depth analysis of iSNVs to identify and characterize the functional properties of non-synonymous mutations found in ORF-1 a/b, S, and N genes. Lastly, we detected a minority variant in non-vaccinated patients in Kenya, with an immune escape mutation S255F of the spike gene and showed differential recombinant RNA species. Overall, this work identified unique in vivo mutations and intrahost recombination patterns in SARS-CoV-2 which could have significant implications for virus evolution, virulence, and immune escape.

## Introduction

Previous studies have implicated both minimal and significant increases in the intrahost diversity of viruses with vaccination ^1–3^. Vaccination is a critical mitigation factor in controlling the COVID-19 pandemic and in Kenya it began with adults in March 2021 and later proceeded to teenagers in November 2021^4^. As of April 2023, 23 million vaccines have been dispensed to approximately 12 million adults and 2 million children (below 18 years). Of the 12 million adults, 10 million were fully vaccinated, whereas 2 million had received one dose of the vaccine ^4^. Nonetheless, by 2023 only 37.2 % of adults and 10.1% of children were vaccinated, which is lower than in other countries globally such as the USA, where 78.9% of adults and 77.4% of children are vaccinated ^4^.

A recent study evaluating the immunity of SARS-CoV-2 with vaccine uptake in Kenya highlighted issues with hesitancy and inequity in society ^5^. It was reported that vaccine hesitancy was mainly due to concerns over safety, efficacy, and religious and cultural beliefs ^5^. However, there is a deficiency in studies investigating the evolution and adaptation of the virus and the host during vaccination, which could highlight broader perspectives on the overall effects of the vaccine and population immunity.

SARS-CoV-2, the etiological agent of COVID-19, is an enveloped single-stranded RNA virus belonging to the genus betacoronavirus, also comprising of SARS-CoV and MERS-CoV ^6,7^. This virus contains 10 open reading frames (ORFs) and four major structural proteins, namely, Spike (S), Membrane (M), Envelope (E), and Nucleocapsid (N) ^6,7^. Based on previous works, the S gene, ORF1a/b, and the N gene single nucleotide variations (SNVs) significantly affect the virus’s infectivity, transmissibility, and overall fitness^7–10^. For example, the spike protein of the omicron variant has approximately 26-35 mutations, which could affect the protein’s ability to bind to ACE-2 ^11–13^. Also, the ORF-1 has the largest number of missense mutations in the *nsp-3* gene, affecting viral replication ^14^.

Evaluation of intrahost SNVs (iSNVs) is a well-established approach to determine viral adaptation ^15,16^, however, identifying the RNA recombination events that may introduce deletions or insertions into the viral genome can be a step further in understanding viral evolution and its transmission dynamics during epidemics ^17^. SARS-CoV-2 is reported to contain both intrahost, and interhost recombination events, and the receptor-binding domain of the virus is a product of interhost recombination events between coronaviruses from pangolins ^18–20^. Numerous SARS-CoV-2 intrahost recombination events and recombinant RNA species are generated by non-homologous recombination and have been identified in cell culture and clinical samples ^17,21,22^. The investigation of intrahost recombinant RNA species, such as sub-genomic RNAs (sgmRNAs), defective RNA genomes (DVGs), micro-Indels, and insertions, is important to understanding virus evolution, adaptation, and the effects on transmission and infectivity ^21–26^. For example, the formation of sgmRNAs is essential for coronavirus replication and are suggested biomarkers for monitoring the progression and infectivity of the virus ^21,26–29^. Micro-insertions and deletions are often detected in the furin cleavage site of the spike protein of SARS-CoV-2 ^23^ and defective RNA genomes have been shown to compete with wild-type viruses changing the fitness of the virus and disease outcomes ^21,25,30^. Therefore, an evaluation of both intrahost SNVs and recombination events could expand our knowledge of the biology behind the infectivity, clinical manifestation, and response to vaccines and therapeutics.

Here we investigate the evolution of SARS-CoV-2 in a cohort of vaccinated and non-vaccinated patients in Kenya. We identify intrahost recombination events in both groups and show similar trends in recombination patterns. We establish that the recombination ‘hotspots’ in both groups are found in the ORF1a/b, S, and N genes. Additionally, we quantify types of recombinant RNA species and show differential expression of sub-genomic RNAs, defective RNA genomes, large insertions, and micro-deletions in vaccinated patients. We also demonstrate the recombination landscape of SARS-CoV-2 between and during transmission waves caused by different variants of concern in Kenya and highlight changes in the production of recombinant RNA species. Further, we resolve unique iSNVs in vaccinated and non-vaccinated patients on the ORF 1a/b, S, and N genes, and characterize their functional properties. Lastly, we reveal a minority variant occurring in non-vaccinated patients, which could have immune escape properties. Overall, this work sheds light on how the genetic evolution of SARS-CoV-2 in the Kenyan population is affected by vaccination and the introduction of new variants, through an in-depth analysis of both intrahost SNVs and intrahost recombination events.

## Results

### A sub-cohort of vaccinated and non-vaccinated samples from COVID-19 patients in Kenya

We collected 1589 nasopharyngeal swabs from patients who tested positive for SARS-CoV-2 via RT-PCR between June 2020 and December 2022, in the Kenyan counties of Bungoma, Busia, Homabay, Kakamega, Kisii, Migori, Nyamira, Trans Nzoia, Vihiga, and West Pokot (Fig. 1A). We extracted total RNA and performed whole genome sequencing of SARS-CoV-2 using ARTIC primer pools to generate cDNA libraries. These libraries were sequenced on the NextSeq and MiSeq Illumina platforms. Sequencing data were processed using nf-core/viralrecon v2.5, yielding ≥90 sequence coverage for each sample. For the vaccination analysis, we selected a sub-cohort of 305 sequences, with ≥ 99% genome coverage, obtained from October 2021 to December 2022, which coincides with the commencement of COVID-19 vaccination in Kenya. This selection minimized clade variability, as most sequences belonged to the omicron clade. This sub-cohort included 187 sequences from non-vaccinated and 118 from vaccinated patients. All samples included information on the vaccination status as either yes or no and had 179 females and 126 males, with ages ranging between 0 to >50 years (Fig 1B). See the baseline characteristics of the vaccination analysis sub-cohort (Table 1).

**Figure 1:**
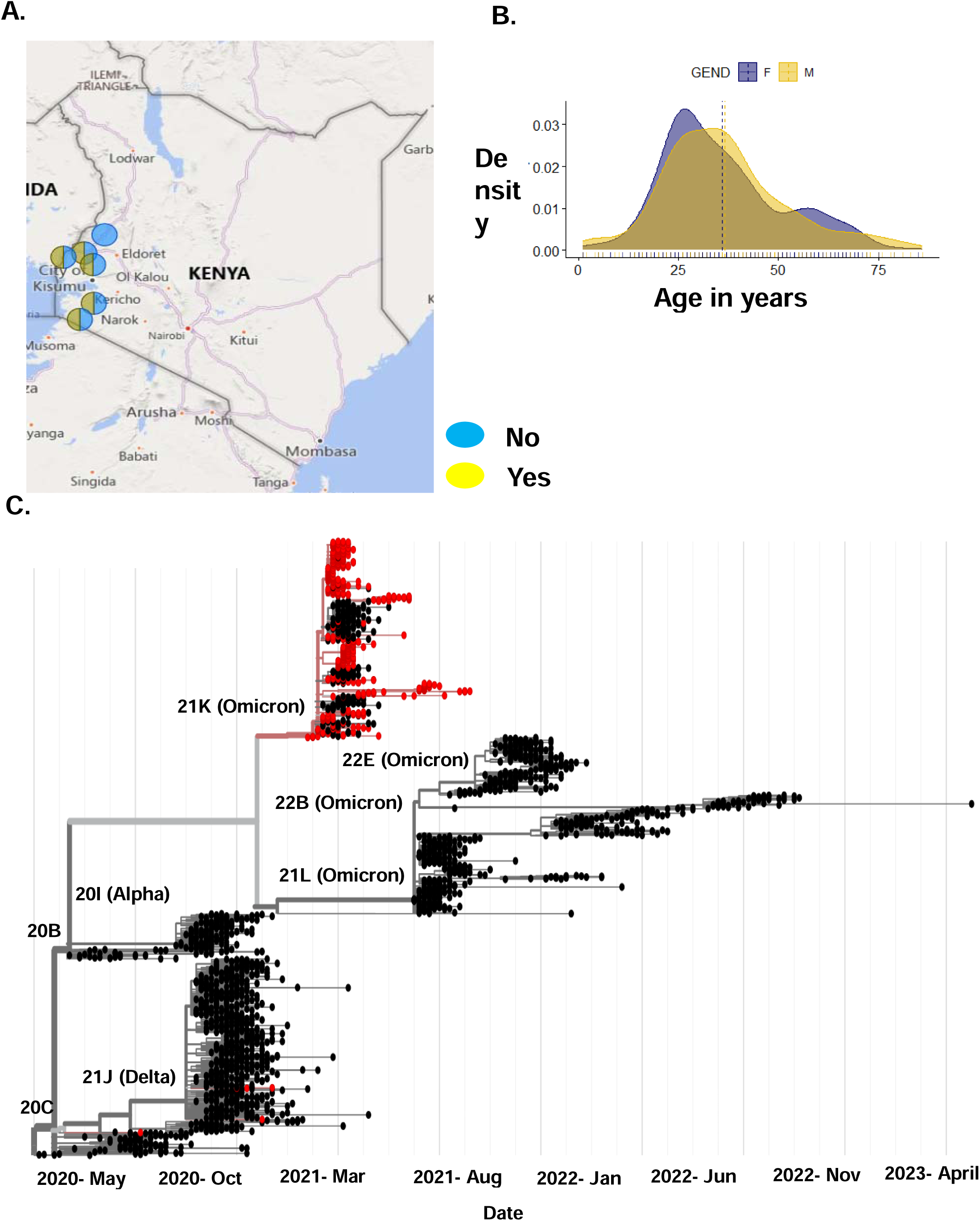
Demographic of vaccinated and non-vaccinated patients in Kenya. A) Shows the locations of the vaccinated and non-vaccinated patients, dark blue circle shows samples with the vaccination status as no and yellow represents the vaccination status as yes. B) Shows the age in years and gender of the cohort. C) Shows the phylogenetic analysis of our non-vaccinated and vaccinated samples based on global trends. Samples in red represent our cohort sequences and samples in black are publicly available sequences.

**Table 1:** Baseline characteristics of vaccinated and non-vaccinated sub-cohort used in this study. The table provides details of the sex, age, Ct values, clade, and vaccine dosage in this cohort.

|  | NON-VACCINATED | VACCINATED | TOTAL |
| --- | --- | --- | --- |
| **<u>SEX</u>** |  |  |  |
| MALE | 84 (45%) | 42 (36%) | 126 (41%) |
| FEMALE | 103 (55%) | 76 (64%) | 179 (59%) |
| **<u>AGE</u>** |  |  |  |
| 0-30 | 73 (39%) | 61 (52%) | 134 (44%) |
| 30-50 | 80 (42%) | 38 (32%) | 118 (39%) |
| >50 | 34 (19%) | 19 (16%) | 53 (17%) |
| **<u>CT VALUE</u>** |  |  |  |
| <35 | 158 (84%) | 82 (69%) | 240 (79%) |
| >35 | 0 (0%) | 0 (0%) | 0 (0%) |
| N/A | 29 (16%) | 36 (31%) | 65 (21%) |
| **<u>CLADE</u>** |  |  |  |
| 20A | 0 (0%) | 1 (1%) | 1 (0.2%) |
| DELTA | 3 (2%) | 2 (2%) | 5 (1.8%) |
| OMICRON | 184 (98%) | 115 (97%) | 299 (98%) |
| **<u>VACCINE DOSAGE</u>** |  |  |  |
| COMPLETE | 145 (78%) | 0 (0%) | 145 (78%) |
| NOT COMPLETE | 36 (19%) | 0 (0%) | 36 (19%) |
| N/A | 5 (3%) | 1. (0%) | 5 (5%) |

We mapped this sub-cohort to globally available sequences on UShER, where we show that the sequences were mainly from Delta and Omicron SARS-CoV-2 variants (Fig. 1C). This was expected based on the timing of sample collection. We identified the most frequently occurring single nucleotide polymorphisms (SNPs) in this cohort and noted that majority are found on the S gene, ORF1a/b, and the N gene (Supplementary Fig. 1). The most frequently occurring mutations in the S gene were D614G, H69_V70 deletion, T95I, G142_Y145 deletion, and T547. For the ORF1a/b gene, T3255I, L4715L, L3674_G3676 deletion, I3758V, and P3395H occurred the most, while in the N gene, P13L, E31_S33del, R203K, and G204R were the most occurring.

### Analysis of intrahost recombination patterns and recombinant RNA species in the vaccinated and non-vaccinated sub-cohort

With an increase in genomic surveillance, SARS-CoV-2 intrahost recombination events of interest have been reported globally, making recombination a key factor in virus evolution ^17,31^. We evaluated intrahost recombination events and the resulting recombinant RNA species to characterize SARS-CoV-2 genetic diversity in our vaccinated and non-vaccinated sub-cohort. Notably, all samples used in this analysis were processed and sequenced using standardized laboratory and bioinformatics methods, ensuring a fair comparison. Furthermore, the majority of the samples belong to the omicron clade, minimizing variation by clade as a confounding factor.

We used ViReMa, a viral recombination mapper that identifies recombination events including deletions, insertions, duplication, copy-back, snap-back, and viral-host chimeric events as described previously ^32–34^. ViReMa requires no prior information on recombination sites, allowing the discovery of previously unknown and/or complex recombination events ^32–34^. We observe similar intrahost recombination patterns between vaccinated and non-vaccinated patients (Fig. 2A, Supplementary Fig. 2). The SARS-CoV-2 genome positions with the highest recombination events were 2883 - 2902, 11286 - 11296, 21986 – 21996, 28362 - 28372, 75 - 27047, 76 - 26480, and 75 – 21526 (Fig. 2A). Of these events, the most common were between 2883 −2902 and 11286-11296 on the ORF1a/b, 21986-21996 on the S gene and 28362-28372 on the N gene. Prior work assessed the recombination of SARS-CoV-2 *in vitro* by looking at top recombinant species and recombination frequency based on genome position^21^. *In vivo* findings align with this work where there was increased recombination at the start and end of the genome and that sub-genomic RNAs and defective viral genomes had the highest counts (Supplementary Fig. 3 A & B).

**Figure 2:**
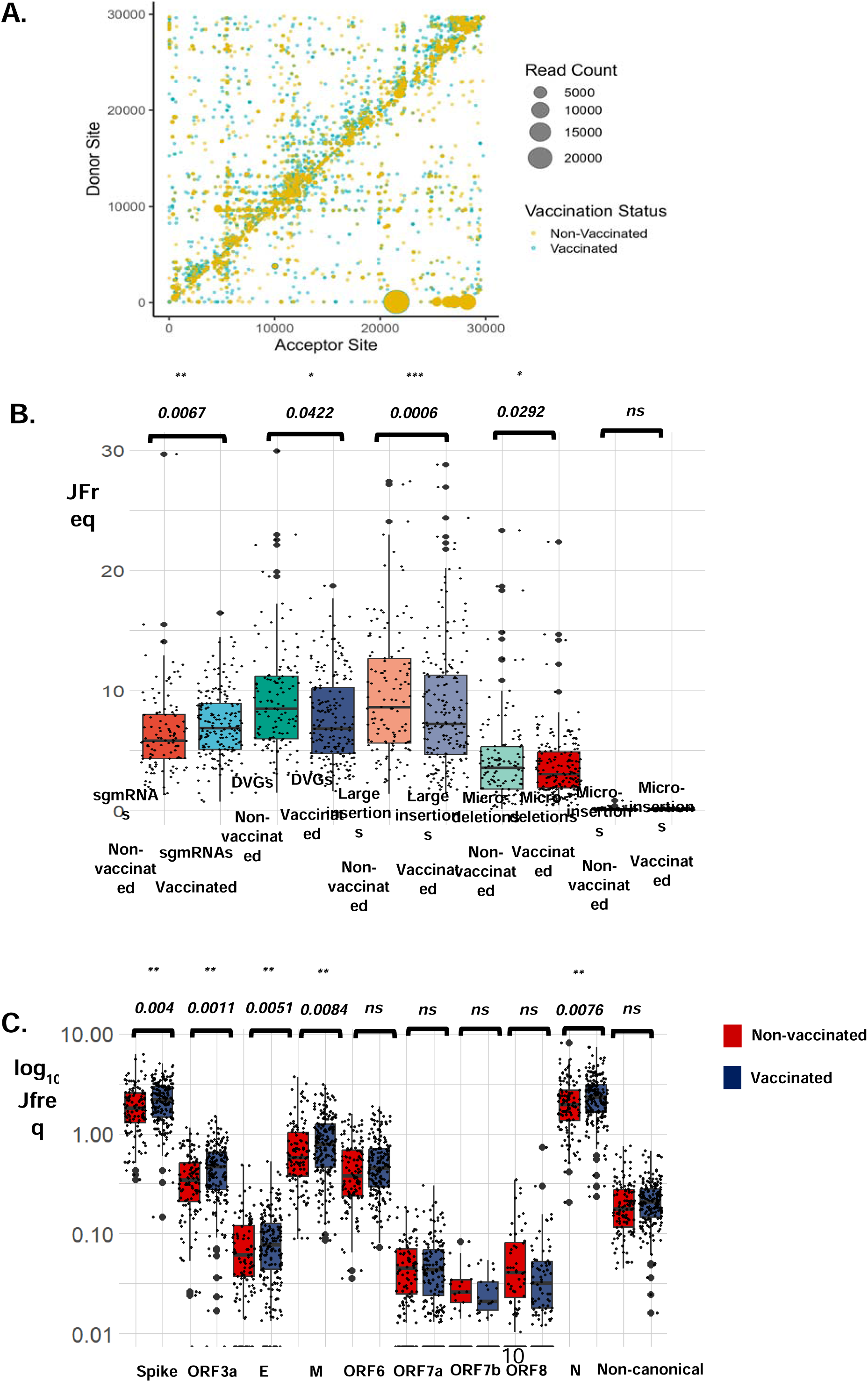
ViReMa identifies recombination events and RNA species in vaccinated and non-vaccinated patients. A) The scatter plots show the mapping of recombination events of the 187 non-vaccinated and 118 vaccinated patients, where the donor site on the y axis is mapped to the acceptor site on the x-axis. The gradient in the legend of the scatter plot represents the number of patient samples containing at least a recombination event. The darker shaded circles in the scatter plot represent events that occur in multiple patient samples, while the circle size corresponds to the count of the reads of a recombination event. B) The Box plot represents the distribution in the JFreq counts of sgmRNAs, DVGs, large insertions, micro-deletions, and micro-insertions between vaccinated and non-vaccinated patients. An unpaired t-test was used to determine statistical significance. C) The Box plots show the types of sgmRNAs found in vaccinated (blue) and non-vaccinated (red) individuals. An unpaired t-test was used to determine statistical significance between each of sgmRNAs in vaccinated and non-vaccinated groups . For the box plots larger quartiles represent where the majority of the data points are represented, the central line shows the median, the whiskers represent the highest and lowest values and the outliers are any data points outside the 1.5x quartile range.

We quantified the recombinant RNA species including sub-genomic RNAs (sgmRNAs), defective viral genomes (DVGs), large insertions, micro-deletions, and micro-insertion events in this cohort (Fig. 2B). We used a junction frequency (JFreq) count to represent the frequency of a recombination event ^21^, which is the number of NGS reads in which a junction is detected by ViReMa normalized to the total reads in a dataset mapping to the viral genome (total mapped reads)^21^. We then performed an unpaired two-tailed student t-test to determine statistical differences between the vaccinated and non-vaccinated groups in each type of RNA species (Supplementary Table 1). Markedly, we detect a significant increase in the number of sgmRNAs (p-value < 0.0067), and a decrease in DVGs (p-value < 0.042), large insertions (p-value < 0.0006), and micro-deletions (p-value < 0.0292), in vaccinated individuals compared to the non-vaccinated (Fig. 2B). We noted no significant changes in micro-insertions between the two groups (Fig. 2B). Further, to determine if other variables such as sex, age groups, and vaccine dosage affected these findings, we repeated this analysis and observed no significant difference between the groups (Supplementary Fig. 4, 5 & 6). However, we observed that sgmRNAs were exceptionally and consistently higher in vaccinated compared to the non-vaccinated samples irrespective of sex or age group (Supplementary Fig. 4 & 5).

Sub-genomic RNA expression is essential to successful viral replication and assembly, through the production of SARS-CoV-2 structural proteins ^35^. We performed a comprehensive analysis of the expression of non-canonical and canonical sgmRNAs, consisting of conserved structural proteins S, E, M, and N, and accessory proteins ORF 3a, ORF 6, ORF 7a, ORF 7b, ORF 8, and ORF 10. The highest expressed sgmRNAs were for the N and S genes (Fig. 2C). Our data is consistent with previous findings on sgmRNA in SARS-CoV-2 where N and S RNA were the most abundantly expressed ^26,36^. Additionally, based on statistical analyses, there were significant differences in expression in S, ORF3a, E, M, and N sgmRNAs between vaccinated and non-vaccinated groups (Fig. 3C). We also determined whether sex and age group affected this analysis and observed no significant differences (Supplementary Fig. 7 & 8). These findings suggest that the S and N gene expression are the primary components driving the observed upregulation of sgmRNAs in the vaccinated cohort (Fig. 2B).

**Figure 3:**
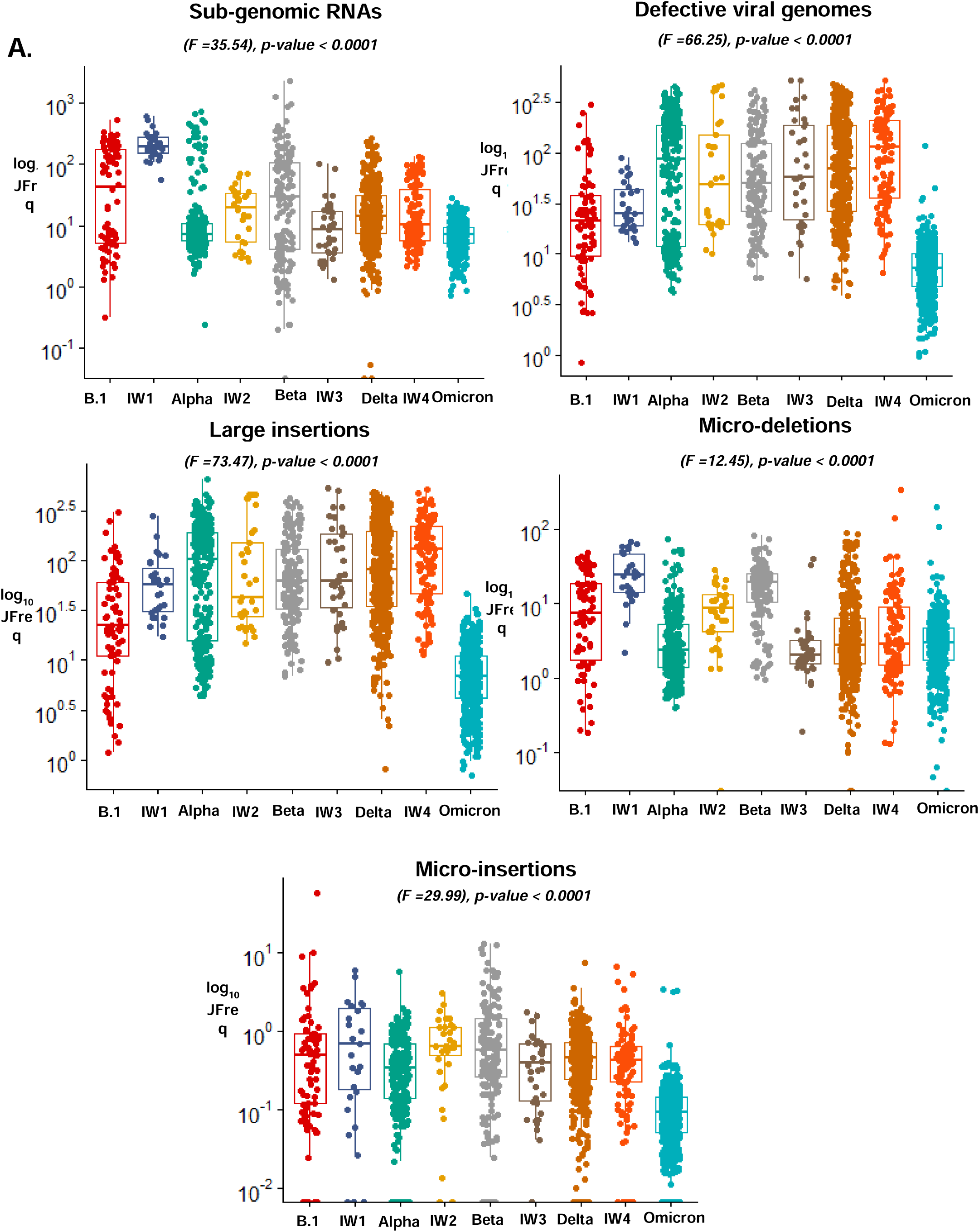
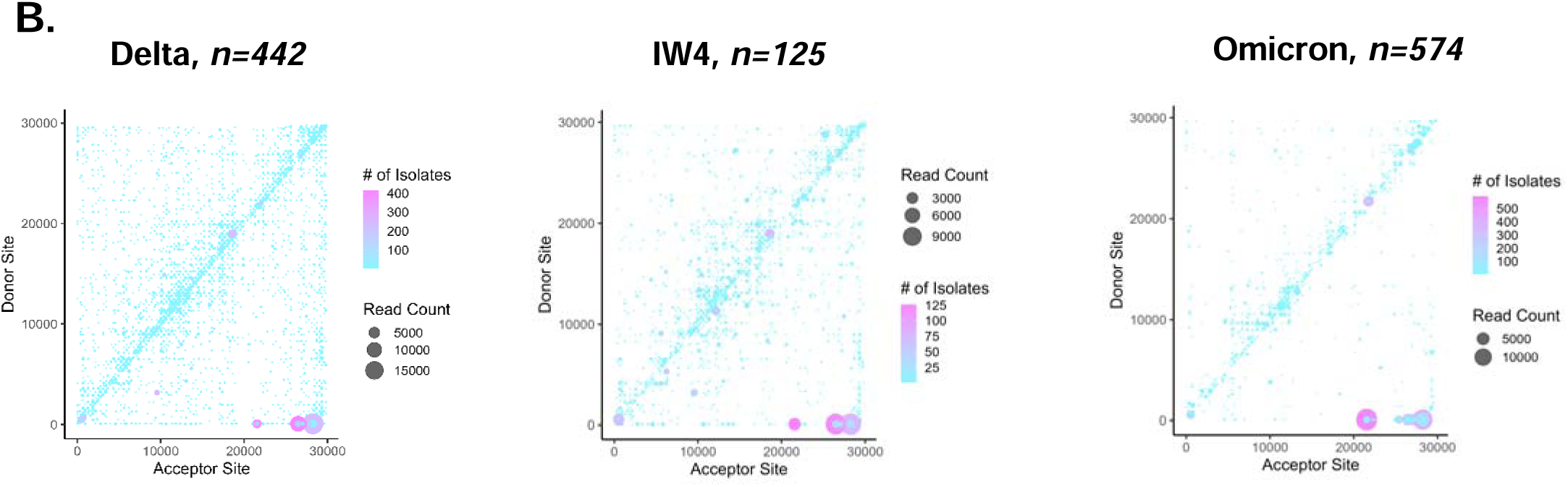
ViReMa identifies recombination events between and during the peak of SARS-CoV-2 transmission waves. A. Shows boxplots of the JFreq (junction frequency) quantification of the recombination RNA species between and within SARS-CoV-2 variant infection waves. Statistical significance was determined using one-way ANOVA and Tukey multiple comparison tests. For the box plots larger quartiles represent where the majority of the data points are represented, the central line shows the median, the whiskers represent the highest and lowest values and the outliers are any data points outside the 1.5x quartile range. B. ViReMa scatter plots of SARS-CoV-2 recombination events and hotspots over Delta, interwave 4, and Omicron variants. The gradient in the scatter plot legend represents the number of patient samples containing a recombination event. The darker shaded circles in the scatter plot represent events that occur in multiple patient samples, while the circle size corresponds to the count of the reads of a recombination event.

### An evaluation of SARS-CoV-2 transmission waves reveals differential recombinant RNA species and patterns

The possibility of inter-variant recombination was assessed. Following the pattern of SARS-CoV-2 transmission waves in Kenya ^37^, we grouped all 1589 sequencing samples from our initial cohort into two categories. Samples collected at the peak of transmission of a particular variant (lineage) and samples collected in the transition period between two variant waves (interwave).

We quantified recombinant RNA species within these groups and used a one-way ANOVA to detect differences between and during the transmission waves. We also performed Tukey multiple comparison tests to detect statistical variances between the groups (Supplementary Table 2). We detected significant changes in the JFreq of sgmRNAs (F=35.54) p-value <-0.0001, DVGs (F=66.25) p-value <0.0001, large insertions (F=73.47) p-value <0.0001, micro-deletions (F=12.45) p-value 0.001, and micro-insertions F(29.99) p-value 0.0001 between all transmission waves (Fig. 3A). The largest variation based on the F score, was in large insertions (F=73.47) and DVGs (F=66.25) (Fig. 3A).

From the Tukey multiple comparison tests, the highest mean differences were observed between sgmRNAs, defective viral genomes, and large insertions (Supplementary Table 2). In sgmRNAs, the largest mean difference was between transmission wave IW1 and Omicron (±223.2), and the lowest was between IW4 and Delta (±0.4706), while for DVGs the highest was between IW4 and Omicron (±129.9), and the lowest between IW2 and IW3 (±0.905). Here, we noted that between sgmRNAs, the highest mean difference were between the start of the pandemic (B.1) and towards the later part of the pandemic (Omicron), suggesting variations in the RNA species regulation during different timepoints of the pandemic. Further, large insertions had the highest mean difference between IW4 and Omicron (±139.2) and the lowest between IW2 and IW3 (±0.5278), whereas micro-insertions and micro-deletions had the highest mean differences between B.1 and Omicron (±1.556) and IW1 and Omicron (±25.37), and the lowest between IW3 and Delta (±0.06) and IW3 and Omicron (±0.1975), respectively (Supplementary Table 2).

Interestingly, we observed that between the transmission waves, some of the most prominent changes were seen during Delta, IW4, and Omicron variants (Fig. 3A) . This was of interest as it coincided with the timeline in which vaccination started in Kenya. We show that DVGs and large insertions had the top most mean differences between Delta and Omicron variants. Specifically, between these waves there was a mean difference of (±110) in DVGs and (±116.7) in large insertions, whereas between IW4 and Omicron had a difference of (±129.9) in DVGs and (±139.2) in large insertions. To ensure changes in genomic coverage didn’t drive these observations, we compared the number of reads and mean depth in these variants and showed no significant differences (Supplementary Fig. 9).

We also assessed recombination hotspots on the SARS-CoV-2 genome between and within the transmission waves and identified the location and frequency of recombination events. We noted an increase in recombination hotspots in most of the interwave periods of transmission compared to the peaks of transmission wave which may point to mixed variant infections (Supplementary Fig. 10). We highlight changes between Delta, IW4, and Omicron variants, and show that in the latter there was a natural selection of recombination events in the ORF1 a/b, S, and N genes (Fig. 3B). Overall, this data reveals insights into the recombination activities within and between peak variants transmission waves.

### Analysis and characterization of intrahost SNVs between non-vaccinated and vaccinated patients reveals unique non-synonymous mutations with key functional properties

Recombination analysis identified genome positions of the four most common deletion events as 2883 −2902 and 11286-11296 on the ORF1a/b, 21986-21996 on the S gene, and 28362-28372 on the N gene. We analyzed the iSNVs occurring in these recombination ‘hotspots’ to gain more insight into the virus genetic evolution. Of all the iSNVs within the recombination hotspots, 67% (215) were non-synonymous, whereas 33% (108) were synonymous.

Using a Venn diagram we demonstrated shared and unique iSNVs between vaccinated and non-vaccinated groups (Fig. 4 A,B & C). We mapped all non-synonymous iSNVs on the S, ORF1a/b, and N genes to determine their distribution on the functional domains of each gene product. As shown on the schematic representations of the gene products, mutations on the S gene were found to be distributed across the entire protein covering all the domains (Fig. 4A), and 6 out of the 8 (75%) were close to the S1/S2 furin cleavage site (FCS) and unique to vaccinated patients (Fig. 4A). The FCS is highly mutable and is a key target by the neutralizing antibodies and binding of the virus particle to the host, which initiates infection, showing the relevance of these changes. ^38–40^. Also of interest, mutation K417Q was found, which lies close to the interaction interface between the Spike protein and ACE-2 receptors of the host ^41,42^. Interestingly, while we observe the K417Q SNV, several previous studies have shown this position to be mutated from Lysine to Threonine ^41,42^.

**Figure 4:**
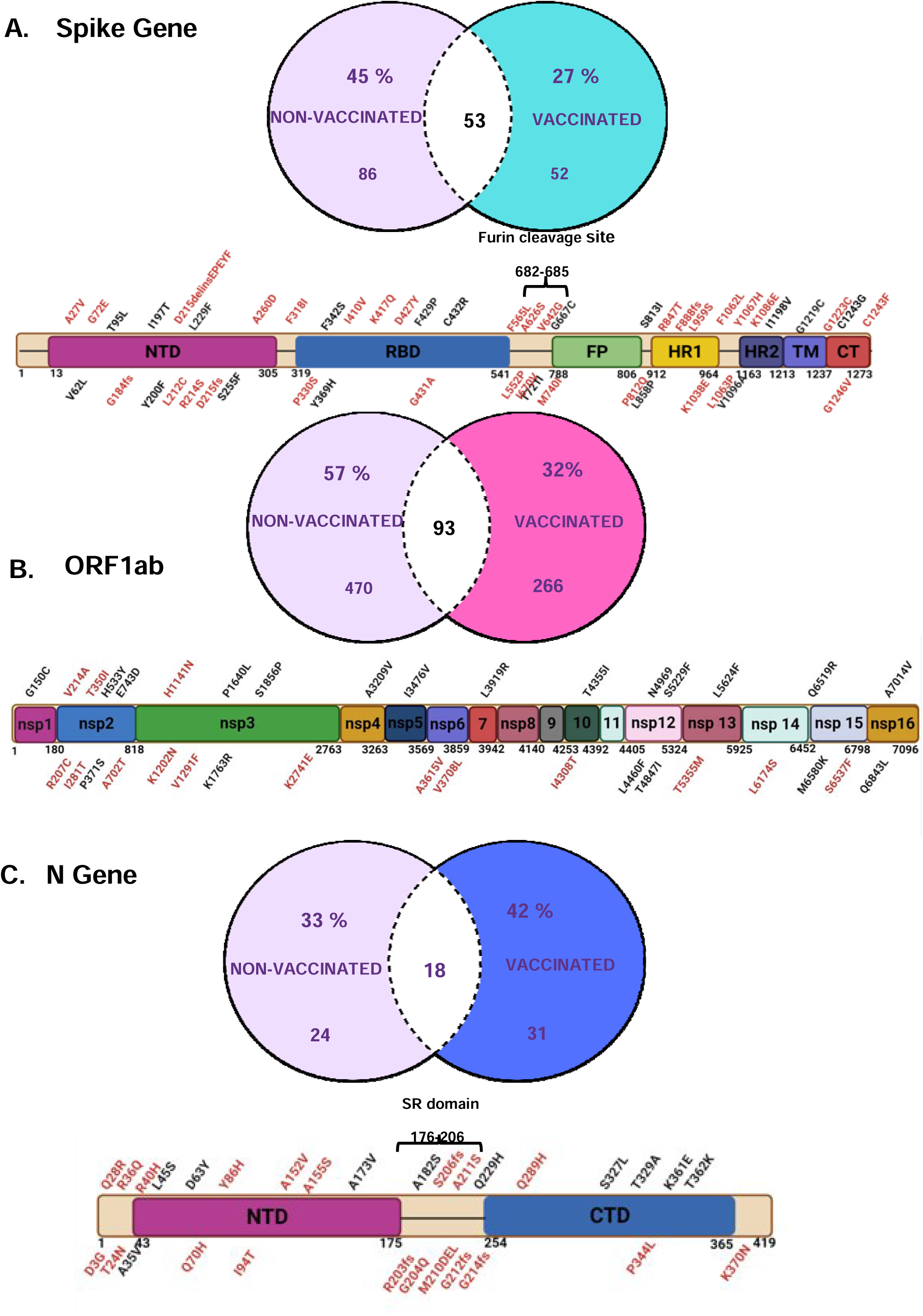
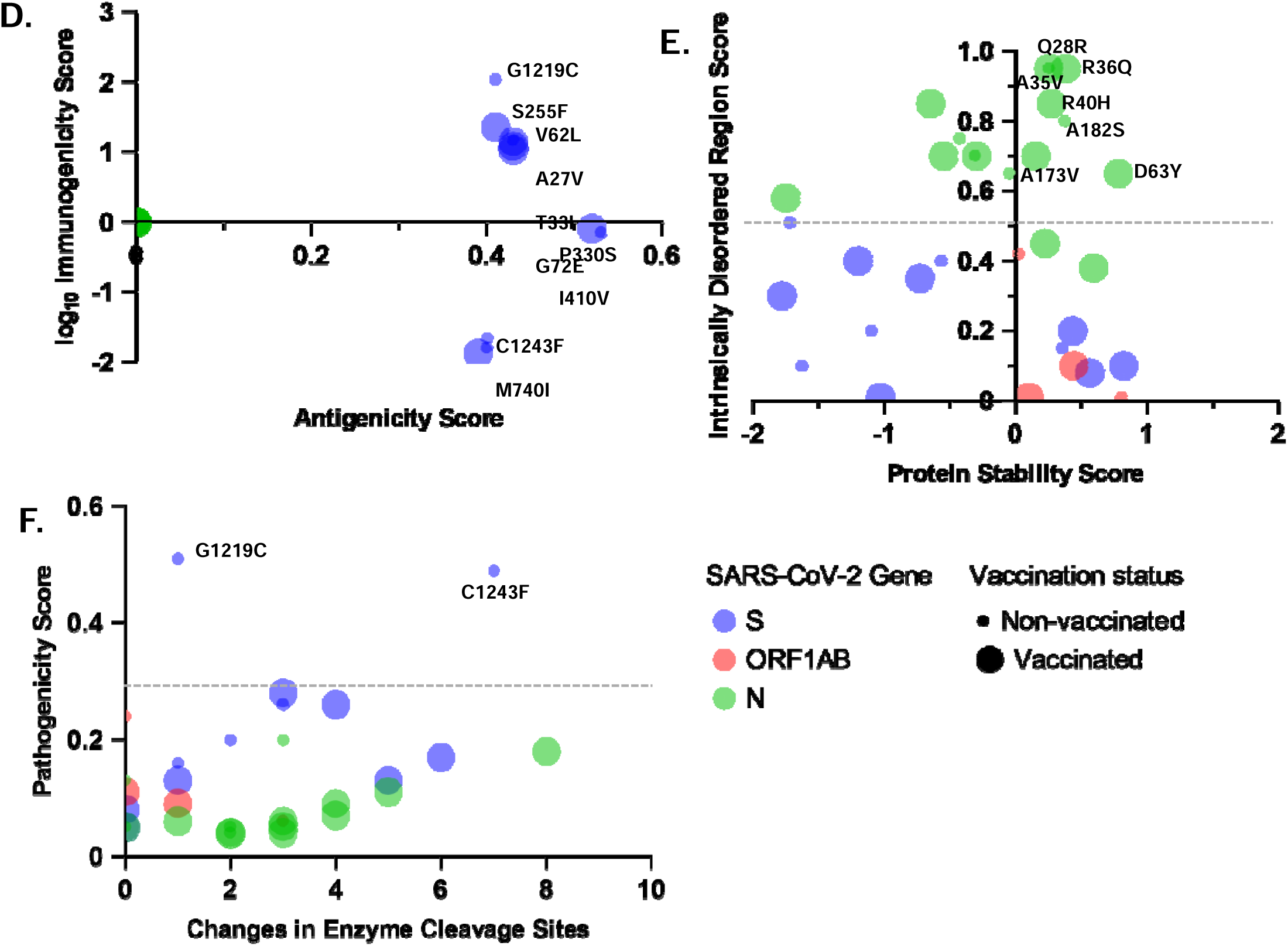
Analysis of unique non-synonymous iSNVs on the S, ORF 1 a/b, and N genes in vaccinated and non-vaccinated patients. A-C. The Venn diagrams show unique and shared iSNVs between vaccinated and non-vaccinated groups. The schematics of the S, ORF1a/b, and N genes show the distribution of unique mutations across the domains of the SARS-CoV-2 gene products. Mutations written in black letters represent those found in non-vaccinated patients and those in red letters represent those in vaccinated patients. D. Shows a multivariable plot between antigenicity scores and log immunogenicity scores of unique mutations. E. Shows a multivariable plot between protein stability score and intrinsically disordered region score of unique mutations. The small dots represent mutations found in the non-vaccinated group and large dots represent mutations found in the vaccinated group. F. Shows a multivariable plot between pathogenicity and no. of changes in enzyme cleavage sites of unique mutations.

Like the S gene, the ORF 1a/b and N unique mutations were distributed across the entire gene product (Fig. 4C). However, we note that in the SR motif of N (between 176-206), 3 out of 4 (75%) mutations are found uniquely in vaccinated patients (Fig. 4C). Specifically, we identify a mutation in position 206 in a vaccinated patient, which has been previously reported as a main target of phosphorylation by kinases^43 44,45^ Phosphorylation of the N gene in the SR domain plays a key role in regulating RNA binding and changing the physical and chemical properties of the N protein ^44,45^.

Further, we accessed the functional impact of the non-synonymous mutations using deep mutation scanning (DMS) datasets, and associated bioinformatics tools. The DMS datasets experimentally characterized all possible mutations on the S gene receptor binding domain (RBD) and the N gene ^46,47^, while COV2Var ^48^ assembled over 13 billion SARS-CoV-2 genome sequences from 2735 viral lineages, 35 different host species, and 218 distinct geographic regions and used various bioinformatics tools to determine the functional properties of 9832 common mutations between all variants^48^.

Of the 215 non-synonymous mutations, 15% (32) passed the threshold of significant changes in protein stability, pathogenicity, location in an intrinsically disordered region, effects on enzyme cleavage, antigenicity, and immunogenicity. We represent these mutations in a multivariable plot that shows the location of the mutation and whether they are found in non-vaccinated (small dots) or vaccinated individuals (large dots) (Fig. 4 D-F).

On the S gene, 33% (4) of the non-synonymous mutations had functional effects on either antigenicity or immunogenicity, based on reference scores on COV2Var (Supplementary Table 3, Table 2). Like other reports, mutation S255F (Fig. 4D) ^49,50^ showed immune escape properties, however, newly identified spike iSNVs of importance were seen. For instance, G1219C causes a substantial increase in pathogenicity and was found in an intrinsically disordered region signifying its role in immune evasion and antibody escape (Fig. 4F). Additionally, not much has been reported on P330S and M740I, which had significant changes in antigenicity and immunogenicity, respectively (Table 2, Supplementary Table 3). Overall, all iSNVs on ORF1a/b had increased protein stability Fig. 4 E), whereas those on the N gene were found in intrinsically disordered regions (Fig. 4F). These findings match previous reports where the N gene mutations in intrinsically disordered regions are linked to immune evasion and antibody escape ^51^.

**Table 2:** Functional characteristics of non-synonymous mutations found in COV2Var database and DMS experiments.

|  |  | individuals<br>with<br>mutation | status | Stability<br>Threshold<br>> 0 | Threshold ><br>0.5 | Disordered<br>Region<br>Threshold<br>> 0.5 | /<br>Decreased<br>Enzyme<br>Cleavage<br>Sites |  |  |
| --- | --- | --- | --- | --- | --- | --- | --- | --- | --- |
| G1219C | S GENE | 3 | Non-vaccinated | -1.72 | 0.507 | 0.51 | 1 | 0.4071 | 2.0441 |
| S255F | S GENE | 5 | Non-vaccinated | -0.73 | 0.169 | 0.35 | 6 | 0.4116 | 1.3571 |
| T732I | S GENE | 1 | Non-vaccinated | 0.43 | 0.13 | 0.2 | 1 | 0.3948 | -1.8668 |
| V62L | S GENE | 1 | Non-vaccinated | -1.78 | 0.132 | 0.3 | 5 | 0.4345 | 1.031 |
| A27V | S GENE | 1 | Vaccinated | 0.82 | 0.084 | 0.1 | 0 | 0.4288 | 1.0841 |
| C1243F | S GENE | 1 | Vaccinated | 0.35 | 0.494 | 0.15 | 7 | 0.404 | -1.7921 |
| G72E | S GENE | 1 | Vaccinated | -1.2 | 0.259 | 0.4 | 4 | 0.4263 | 1.1465 |
| I410V | S GENE | 1 | Vaccinated | -1.03 | 0.05 | 0.01 | 0 | 0.5231 | -0.0868 |
| M740I | S GENE | 1 | Vaccinated | -1.1 | 0.204 | 0.2 | 2 | 0.4017 | -1.658 |
| P330S | S GENE | 1 | Vaccinated | -1.63 | 0.255 | 0.1 | 3 | 0.5318 | -0.1477 |
| Q628K | S GENE | 1 | Vaccinated | 0.56 | 0.277 | 0.08 | 3 | 0 | 0 |
| T33I | S GENE | 1 | Vaccinated | -0.57 | 0.156 | 0.4 | 1 | 0.4299 | 1.1738 |
| A3209V | ORF1AB | 3 | Non-vaccinated | 0.8 | 0.24 | 0.01 | 0 | 0 | 0 |
| A3615V | ORF1AB | 4 | Vaccinated | 0.1 | 0.105 | 0.01 | 0 | 0 | 0 |
| K1202N | ORF1AB | 2 | Vaccinated | 0.02 | 0.057 | 0.42 | 3 | 0 | 0 |
| T350I | ORF1AB | 2 | Vaccinated | 0.44 | 0.093 | 0.1 | 1 | 0 | 0 |
| A173V | N GENE | 1 | Non-vaccinated | 0.15 | 0.049 | 0.7 | 0 | 0 | 0 |
| A182S | N GENE | 1 | Non-vaccinated | 0.37 | 0.036 | 0.8 | 2 | 0 | 0 |
| T362K | N GENE | 1 | Non-vaccinated | -1.75 | 0.087 | 0.58 | 4 | 0 | 0 |
| S327L | N GENE | 1 | Non-vaccinated | 0.59 | 0.113 | 0.38 | 5 | 0 | 0 |
| Q229H | N GENE | 1 | Non-vaccinated | 0.22 | 0.055 | 0.45 | 1 | 0 | 0 |
| D63Y | N GENE | 1 | Non-vaccinated | 0.78 | 0.181 | 0.65 | 8 | 0 | 0 |
| A35V | N GENE | 1 | Non-vaccinated | 0.25 | 0.045 | 0.95 | 0 | 0 | 0 |
| 6 | N GENE | 1 | Vaccinated | 0.25 | 0.047 | 0.95 | 3 | 0 | 0 |
| Q289H | N GENE | 1 | Vaccinated | -0.43 | 0.202 | 0.75 | 3 | 0 | 0 |
| A211S | N GENE | 1 | Vaccinated | -0.55 | 0.035 | 0.7 | 2 | 0 | 0 |
| A152V | N GENE | 1 | Vaccinated | -0.05 | 0.13 | 0.65 | 0 | 0 | 0 |
| R36Q | N GENE | 1 | Vaccinated | 0.38 | 0.061 | 0.95 | 3 | 0 | 0 |
| T24N | N GENE | 1 | Vaccinated | -0.65 | 0.042 | 0.85 | 2 | 0 | 0 |
| K370N | N GENE | 1 | Vaccinated | -0.3 | 0.04 | 0.7 | 3 | 0 | 0 |
| A155S | N GENE | 1 | Vaccinated | -0.31 | 0.045 | 0.7 | 2 | 0 | 0 |
| R40H | N GENE | 1 | Vaccinated | 0.27 | 0.067 | 0.85 | 4 | 0 | 0 |

We conducted a hypergeometric test to determine the likelihood of success of antigenicity, immunogenicity, and protein function for each iSNV compared to other COV2Var mutations. Analysis of our sample revealed that most of the iSNVs were underrepresented in terms of antigenicity, immunogenicity, and protein stability. Specifically, there was a 10-fold underrepresentation in antigenicity P(X=x) = 0, an 8-fold underrepresentation in immunogenicity P(X=x) = 0.00031, a 3-fold increase in SNVs that increase protein stability P(X=x) = 0, and an 11-fold decrease in SNVs reducing protein stability P(X=x) = 0.(Supplementary Table 3).

### Evaluation of a minority variant with linked co-mutations and recombination events

We sought to determine if the unique iSNV mutations, based on vaccination status, in the S, N, and ORF1a/b genes occur in the same patient and if there was any correlation with recombination events. We evaluated the mutations based on the location of the patients, the number of patients in the cohort, and the frequency of recurring (Table 3). In the S gene of the non-vaccinated group, samples from Bungoma, Kakamega, Kisii, Migori, and Nyamira counties showed common mutations that are unique to non-vaccinated patients (Table 3). The most frequent mutation in the S genes was S255F, found in 5 out of 15 patients in Nyamira county. We also identified mutation G1219C in 2 patients in Migori County. On the ORF1a/b genes, unique mutations were found in Bungoma, Kakamega, Migori, and Nyamira counties. The most frequently occurring mutation in ORF1a/b was I3476V, found in 10 out of 15 (66.7%) patients in Nyamira. Other mutations frequently occurring in Nyamira samples were N4969S (5/15; 33.33%), S5229F (5/15; 33.33%), and P1640L (4/15; 26.67%) (Table 3).

**Table 3:**
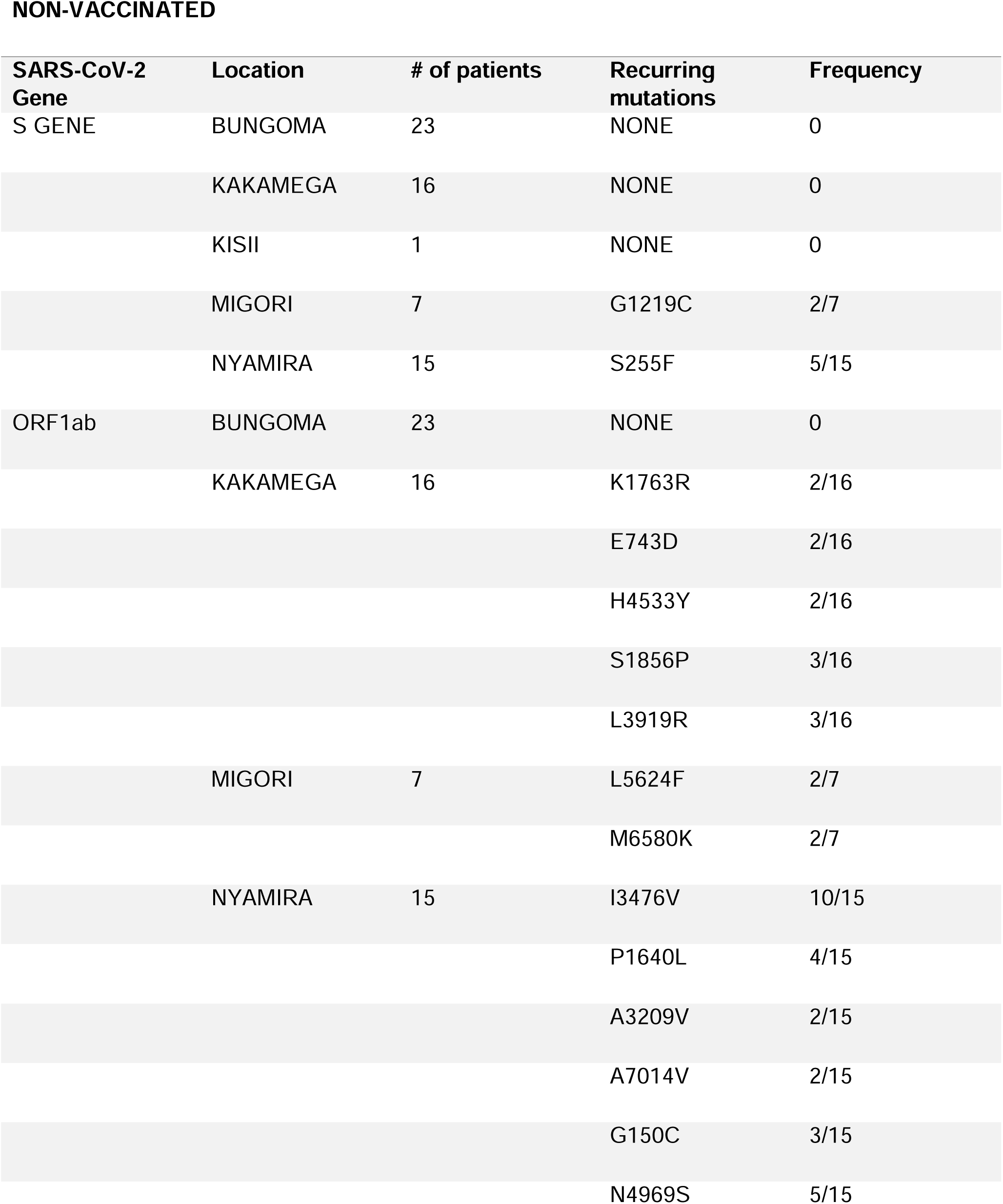
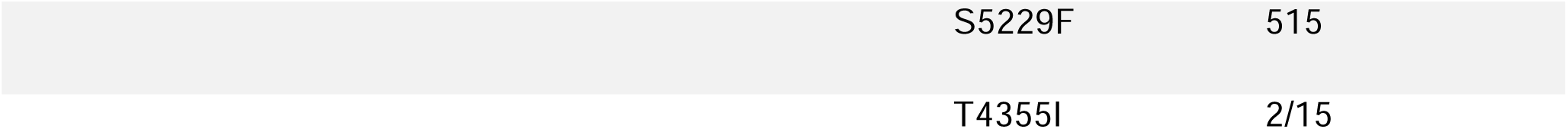
Patients with unique mutations in non-vaccinated patients in Kenya.

Mutations unique to the vaccinated group of patients sampled from Bungoma, Kakamega, Migori, and Busia were also identified (Table 4). In the S gene, L212C (2/4; 50%) and R214S (2/4; 50%) were the most frequently occurring mutations in Kakamega. Whereas on the ORF1a/b gene, V2149A (6/19; 31.5%) was the most frequently occurring mutation event in Migori, followed by A3615V (4/19; 21%), V3708L (4/19; 21%), and V702T (4/19; 21%).

**Table 4:** Patients with unique mutations in vaccinated patients in Kenya.

VACCINATED
| SARS-CoV-2 Gene | Location | # of patients | Recurring mutations | Frequency |
| --- | --- | --- | --- | --- |
| S GENE | BUNGOMA | 12 | NONE | 0 |
|  | KAKAMEGA | 9 | L212C | 2/9 |
|  |  |  | R214S | 2/9 |
|  | MIGORI | 19 | R346N | 2/19 |
|  | BUSIA | 2 | NONE | 0 |
| ORF1ab | BUNGOMA | 12 | I4308T | 2/12 |
|  | KAKAMEGA | 9 | K2741E | 2/9 |
|  |  |  | T350I | 2/9 |
|  |  |  | K1202N | 2/9 |
|  |  |  | L6174S | 3/9 |
|  | MIGORI | 19 | A3615V | 4/19 |
|  |  |  | V2149A | 6/19 |
|  |  |  | V3708L | 4/19 |
|  |  |  | V702T | 4/19 |
|  | BUSIA | 2 | H1141N | 2/2 |

We next determined if these low-frequently unique iSNVs in the S gene and the ORF1a/b co-occur in the same patients. Interestingly, we observed that in the non-vaccinated cohort, five samples in Nyamira had the same set of unique (only in non-vaccinated patients) mutations on S gene and the ORF1a/b (Fig. 5A). All five patients had mutations S255F on the S gene and I3476V, N4969S, and S5339F on the ORF1a/b. S255F is an important mutation previously identified in the S gene, with immune escape properties ^1,52^, however, the I3476V, N4969S, and S5339F are all unique to the non-vaccinated patients and have not been reported previously (Fig. 5A). This finding suggests a possible spread of a variant (minority variant) within a pocket of population and with an important immune evasion capability based on the presence of S255F.

**Figure 5:**
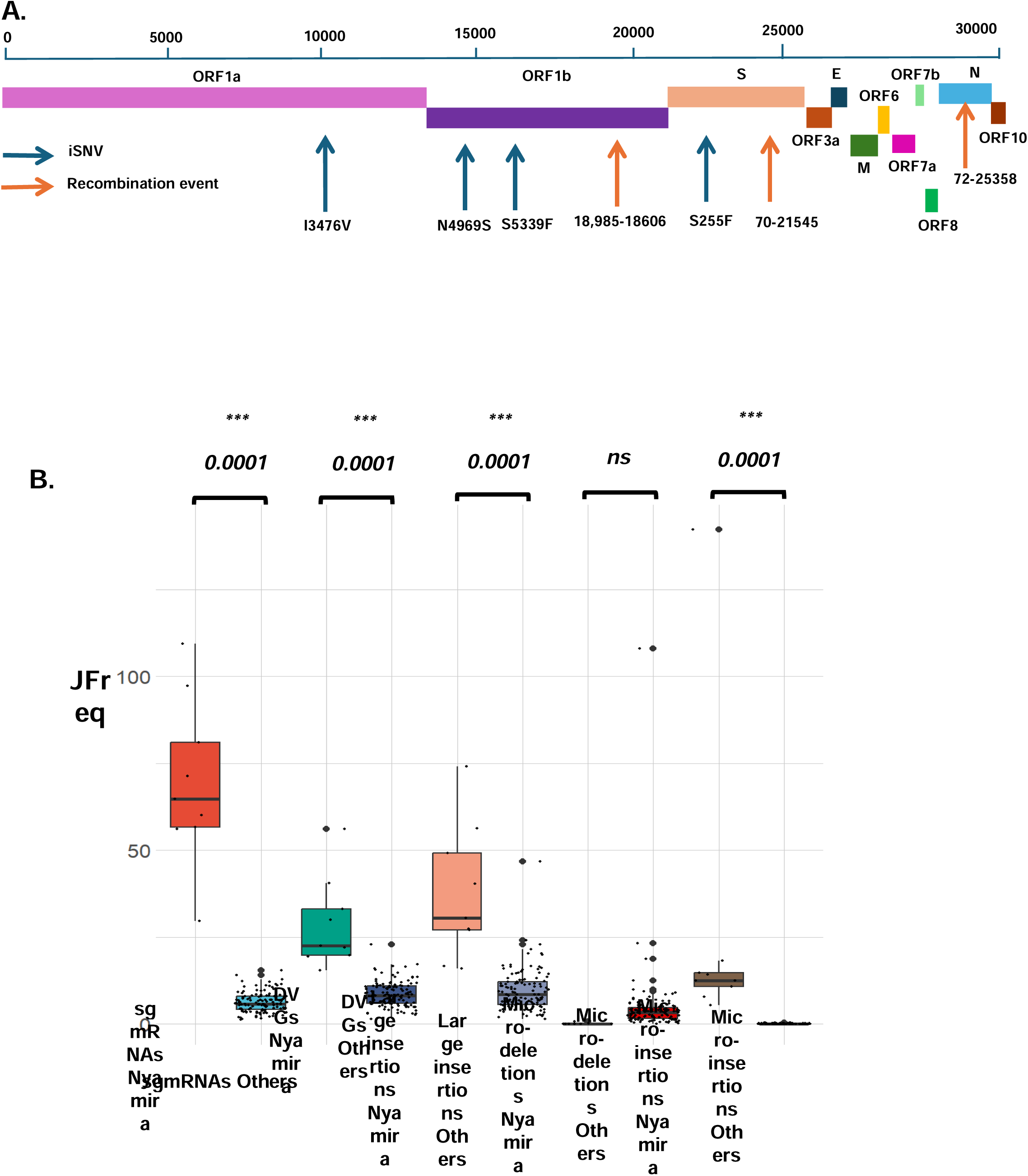
Analysis of the virus diversity of vaccinated and non-vaccinated patients reveals a minority variant. A) Schematic of the SARS-CoV-2 genome, highlighting co-occurring mutations on the S and ORF1a/b and top recombination events in patients in Nyamira county, Kenya. B) Shows boxplots of the JFreq (junction frequency) quantification of the recombination RNA species between the Nyamira patients (n=10) and the other non-vaccinated patients (n=108). Statistical significance was determined using one-way ANOVA and Tukey multiple comparison tests. For the box plots larger quartiles represent where the majority of the data points are represented, the central line shows the median, the whiskers represent the highest and lowest values and the outliers are any data points outside the 1.5x quartile range.

To further characterize this minority variant, we quantified the recombinant RNA species in this group of Nyamira samples and compared them to the rest of other non-vaccinated patients (Fig. 5B). We observed significant increases in the JFreq of sgmRNAs, DVGs, large insertions and micro-insertions, with more than a ten-fold increase in the abundance of sgmRNAs in these individuals (Fig. 5B). This observation suggests that patients with this minority variant had both unique iSNVs and significant changes in the expression of recombinant RNA species, that could affect the virus’s adaptation, fitness, and infectivity. Further work is needed to determine any functional links between these unique iSNVs identified and their role in viral recombination events.

## Discussion

The low uptake of the COVID-19 vaccine in Kenya as with many other African countries could be attributed to the lack of access, mistrust in vaccine efficiency and safety, and religious beliefs ^5^. The low vaccination rate and widespread infection-associated population immunity have limited studies seeking to understand the evolution and diversity of SARS-CoV-2 virus with respect to vaccine-induced immune pressure in the region. Here, we used next-generation sequencing to uncover the diversity and genetic evolution of SARS-CoV-2 through analysis of iSNVs and recombination events related to vaccination status in a cohort within the Kenyan population. Globally, recombination events have been reported in areas with high genomic surveillance, such as the UK, USA, and Denmark ^17,18,31,53–55^, and it is estimated that 5% of circulating US and UK SARS-CoV-2 viruses are recombinant ^16,17^. Specifically, the genomic surveillance of SARS-CoV-2 and the tracking of both intervariant and intrahost recombination events has proven crucial in obtaining a better picture of the virus’ genetic evolution that may be driven by multiple variant infection, immune pressure, and vaccine efficacy ^56^. This data is critical in designing future vaccines, antivirals, and control measures ^16,17,31^.

With respect to vaccination status, we observed large number of recombination events in both vaccinated and non-vaccinated individuals, which may correspond to the general high mutation rate of SARS-CoV-2 virus ^13,39^. We identify similar recombination ‘hotspots’ on the SARS-CoV-2 genome in vaccinated and non-vaccinated patient samples and show that the most common recombination events are found in the ORF1a/b, S, and N genes. These findings corroborate previous studies showing that recombination events occur disproportionately in the spike protein region and that the ORF1a/b gene experiences the largest number of mutations, showing significant virus diversity in these regions ^52,57^.

Coronaviruses lack canonical sites for nonhomologous recombination and possess a high recombination rate, which can result in unpredictable recombination under evolutionary pressures ^17,27,29,33,58^. Here we identify noteworthy increases in sgmRNAs in vaccinated compared to non-vaccinated individuals. This observation is of special interest given the role of sgmRNAs in the expression of viral structural proteins, modulation of host cell translation, and viral evolution ^25,26,59^. Various reports have shown that an increase in sgmRNA affects the ratio and RNA-RNA interactions between sgmRNAs and genomic RNA (gRNA) of SARS-CoV-2, which in turn affects the regulation of discontinuous transcription ^26,60^ Also, the presence of nsp-15 cleavage site in the TRS motif has been linked to a negative feedback mechanism in the regulation of SARS-CoV-2 transcription and replication^60,61^. Additionally, sgmRNAs interact with cellular host factors such as host ribosome MRP RNA, leading to viral degradation ^62,63^.

We also observed a decrease in DVGs in vaccinated individuals within and between transmission waves. DVGs have been shown to interfere with replication of the wildtype virus and can also promote viral persistence in natural infection in RNA viruses such as Hepatitis C and Ebola ^64^. It will be interesting to show if the observed increase in DVG in non-vaccinated patients may be associated with viral persistence in SARS-CoV-2 natural infection. Additionally, SARS-CoV-2 DVGs have robust antiviral efficacy, due to their “higher cost” of evolution, making them good candidates for vaccines ^65,66^. More work is needed to unravel the role of recombinant RNA species in SARS-CoV-2 infection and their mechanism of action in virus evolution and pathogenicity.

We mapped the recombination landscape and recombinant RNA species of SARS-CoV-2 between and within the peaks of variant transmission waves in Kenya^37^. We observed a remarkable difference in frequencies of recombinant RNA species of the virus between transmission waves compared to vaccination. This suggests that as the virus evolves to create new variants the frequency of different recombinant species is altered as a form of adaptation. For example, we observe contrasting differences between the frequency of sgmRNAs during the start of the pandemic (B.1) compared to recent variants in (Omicron). Also, similar to current data, we show that the evolution of SARS-CoV-2 from B.1 to Omicron variants may have led to the natural selection of ORF1 a/b, S, and N genes as recombination hotspots. Several studies have highlighted the importance of these genes in the adaptability, transmissibility, and clinical outcomes of the Omicron variant infections ^11,13,54,67,68^.

Globally, studies have identified types of iSNVs occurring in variants of concern and variants of interest and how these mutations can affect the fitness of the virus and response to vaccines and anti-viral ^8,10,50,52,54,57^. The iSNVs analysis gives insight into SARS-CoV-2 genomic positions and protein domains targeted by virus genetic evolution and their possible association with virus virulence. For example, mutations such as S255F ^1,50^, confer reduced neutralization by monoclonal antibodies and have the potential for immune escape ^1,50^. In this study we identify unique iSNVs, in the context of vaccination status. Some mutations of interest include S255F, which occurred in a group of patients who had previously unreported iSNVs in their ORF1a/b and showed a different pattern of recombination events compared to other non-vaccinated individuals. We use deep mutation scanning experiments and bioinformatics tools to characterize these unique non-synonymous iSNVs showing their relevance in protein structure, pathogenicity, immunogenicity, and effects on affinity between the spike RBD and ACE-2.

Of more interest is the detection of a minority variant in a small pocket of non-vaccinated population that seeks further studies and epidemiological follow-up. These individuals from Nyamira county in Kenya appeared to have S255F mutation in the spike gene and new unreported mutations I3476V, N4969S, and S5339F on ORF1a/b. Although S255F has been previously reported in the literature to cause immune evasion, within an epidemiological cohort, its co-occurrence frequency with the other identified iSNVs in this population is unique and may be a pointer to functional adaptational mechanisms of the virus. We also show that within this same cohort, substantial variations in frequencies of recombinant RNA species in the Nyamira individuals compared to the other non-vaccinated samples are observed. These unique recombination patterns differentiate this cohort of samples from the rest of the population. This case study highlights the significance of using iSNVs and recombination analysis to understand viral genetic evolution and diversity, emphasizing the need for ongoing genomic surveillance across different geographic regions.

The vaccine inequity early in the COVID-19 pandemic phase (early to late 2021) meant that vaccines became widely available in sub-Saharan Africa (SSA) at approximately the same time the Omicron variant occurred (around Oct-Nov 2021). As studies demonstrated, by early 2022, population immunity was >70% across most urban and rural communities in SSA, making it difficult as shown in our studies, to discern the impact of vaccination in SARS-CoV-2 evolution between and during all the different transmission waves ^5^. Additionally, epidemiological data from these studies showed that both vaccinated and non-vaccinated patients had comparable immunity and thus exerted similar immunologic pressures on circulating SARS-CoV2 strains ^5^. While we observed comparable types and frequencies of recombination among vaccinated and non-vaccinated patients, the overall findings present a unique impact of immunologic pressure on the virus. Our data suggest that the delayed vaccination likely induced minimal antival immune pressure capable of driving genetic evolution. Instead, these findings reflect the impact of the combined effect of vaccination and widespread natural-infection-associated immunity in Africa.

Recently, the role vaccination played in within-host selection and population evolution has been of interest ^56^. Using experimental outcomes and mathematical models from similar highly mutating RNA viruses like Influenza, inferences can be drawn on the evolution of SARS-CoV-2^56^. In future studies, information from SARS-CoV-2 evolution studies and epidemiological findings could be used in mathematical models to develop better vaccines and antivirals. For example, non-synonymous mutations, such as those identified in our study, and findings on sub-genomic RNA and defective viral genomes could be used in mathematical models with other epidemiological factors such age, vaccine efficacy, comorbidities, and genetic factors to develop better therapeutics. These findings are bound to change with different regions, for instance, in Africa where the transmission waves of SARS-CoV-2 were experienced with relatively lower levels of vaccination affecting the substitution rates in epitopes (*V*), hence the need for local genomic surveillance and evolution analyses.

This study’s limitations include using a convenient population cohort from the peak of the pandemic and vaccination efforts in Kenya. This sequencing data lacks detailed information on infection history, as well as no negative and contamination controls. Additionally, a lack of comprehensive immune profiling makes it challenging to fully access the impact of vaccination while accounting for pre-existing immunity. Future studies incorporating metadata on infection history and longitudinal analyses could provide a more nuanced understanding of vaccine effects. Despite these limitations, this study offers valuable insights into the intrahost diversity of a vaccinated and non-vaccinated cohort in Kenya, which remains unexplored. Another limitation is the use of ARTIC data, which provides a key limitation in the analysis of recombination, as this data can be noisy due to PCR artifacts. Conversely, to ensure uniformity and accuracy in the analysis, all samples were processed, sequenced, and analyzed in the same laboratory with similar conditions, protocols, and bioinformatics pipelines. This reduced processing and technical variability allowing conclusions from the data to be likely due to biological changes. Also, we compared our ARTIC data to Tiled-ClickSeq data^69^, which was designed and tested to be more sensitive to detecting recombination events and found similarities in the findings. Ultimately, this study underscores the need for increased genomic surveillance in Africa, which will facilitate more research on virus evolution. Such surveillance ensures we can detect drifts in evolution allowing information for updates in vaccines, policy making, and containment of future variants of SARS-CoV-2. In conclusion, the current study shows a broad picture of the differential virus intrahost genetic evolution and diversity between vaccinated and non-vaccinated patients and suggests increased recombination events and hotspots driven mostly by interaction between variants compared to the COVID-19 vaccines. Analysis of recombination events during peaks of transmission waves and the interwaves is a powerful approach to studying virus genetic evolution and its drivers. The work also demonstrates a methodology for studying genetic changes in a pathogen by a simultaneous analysis of both intrahost single nucleotide variations and recombination events.

## Materials and Methods

### Sample collection

We performed sample collection, processing, and analysis in accordance with the Ministry of Health-Kenya COVID-19 pandemic surveillance protocols and guidelines. The study samples were collected over the year 2020 to 2022 and comprised of nasopharyngeal and oral swabs. The samples were kept in viral transport media (VTM) tubes and transported under refrigerated conditions to the ILRI genomics laboratories in three tier packaging systems for processing. An aliquot of 300 microliters was used for RNA extraction, and the remaining was archived in the ILRI’s AZIZI Biorepository.

### SARS-CoV-2 RNA extraction

RNA extraction and purification was performed using the Tan Bead Nucleic RNA extraction kit (Opti Pure Viral Auto tube/plate) (Taiwan Advanced Nanotech Inc. Taoyuan City, Taiwan) following the manufacturer’s instructions. RT-qPCR was performed to identify SARS CoV-2 positive samples using Applied Biosystems Quant Studio 5 Real-Time PCR System (Thermos Fisher Scientific, USA).

### SARS-CoV-2 COVID-Seq Illumina library preparation and sequencing

RT-qPCR positive samples were then selected based on a CT <35 and transitioned for library preparation. Purified RNA was used as template to prepare complementary DNA (cDNA) using random hexamers in a two-step reverse transcriptase process (Illumina COVID-Seq Ruo Kits, Illumina, Inc, USA)^70,71^ . This was followed by tilling/amplification of cDNA using the multiplex ARTIC primer-pools CPP1 and CPP2 version 3 followed by illumina library preparation protocol that uses enrichment bead-linked transposons (EBLT) for fragmentation, size selection, adaptor ligation and PCR enrichment (Illumina COVID-Seq Ruo Kits, Illumina, Inc, USA). The libraries were normalized and pooled to 4 nM before a further dilution to 1.5 pM for loading in NextSeq, or 12 pM for loading in MiSeq illumina sequencing platforms (Illumina, CA, USA) to sequence with the V2 paired-end chemistry^70,71^.

### Variant Calling and Consensus Genome construction

The demultiplexed FASTQ files were merged for every sample and analyzed. Variant calling, and lineage assignment was performed using nf-core/viralrecon v2.5 - a Nextflow-based pipeline ^72^. Briefly, FASTQ files were quality filtered and adapter trimmed using FASTP v0.23.2 with a Phred Score cut-off of 20 ^73,74^. Bowtie2 v2.4.4 was used to map the reads to the reference genome (NC_045512.2) and iVar v1.3.1 to soft-mask primer sequences and identify the variants ^75,76^, with a minimum threshold(-t) 0.25, minimum quality score (-q) 20, and minimum read depthe (-m) 10. SnpEff v5.06 and SnpSift v4.3 were used to annotate and filter relevant mutations identified ^77,78^. Re-construction of the consensus was done with bcftools v.1.15.1.

### ViReMa recombination analysis

Virus-Recombination-Mapper (ViReMa; v0.30 was used to identify and quantify recombination events (insertions, deletion, and duplication) ^32–34^. We used paired-end next-generation sequence data to detect recombination junction events in the SARS-CoV-2 reference genome. For the vaccination data analysis, we retained samples with a genome coverage of >99% coverage and retained 118 non-vaccinated and 187 vaccinated patient samples, and for the transmission waves, we retained samples with genome coverage of >90%. Scatter plots showing ViReMa results were generated on the ViReMaShiny ap and R ^33^ . Scatter plots for both vaccination status and transmission waves included recombination events with a count number of >10. We provide a detailed description of the python script as follows:

#### Transpose_to_WA-1_Coords.py

During the reconstruction of a consensus viral genome from D/RNAseq data (such as ARTIC or Tiled-ClickSeq data) small InDels are fixed by default by pilon. As a result, the coordinates downstream of any corrected InDel are offset by the size of the inserted/deleted nucleotides in the new reference consensus genome. To cross-compare specific recombination events (e.g. such as sgmRNAs corresponding to the structural proteins) between multiple samples, a common coordinate system must be used. ‘Transpose_to_WA-1_Coords’ takes into account the InDels corrected by pilon in the new reference genome, and transposes the reported recombination junctions in the BED file accordingly.

#### Combine_unstranded_annotations.py

This script is appropriate when using D/RNAseq approaches that are unstranded such as, for example, ARTIC Amplicon sequencing protocols. ViReMa generates BED files annotating the genomic coordinates of recombination junctions found during read alignment which contain additional custom columns (when using the -BED12 option) that report the read depth at both the 5’ and 3’ sites of the recombination junction. This scripts takes this BED file as input, and combines the read count at each of these sites if the recombination junctions is found on both negative and positive sense directions (relative to the provided reference genome). To reflect this ambiguity, the new BED file (annotated here as ‘noDir’) reports the junction direction as ‘+/−‘. Due to microhomology commonly found at the donor and acceptor sites of recombination junctions, there is inherent ambiguity as to the exact coordinates of the recombination junction. To address this, ViReMa provides a parameter (‘defuzz’) that ensures that recombination events are annotated at either the 3’-most, 5’-most, or center of the micro-homologous (‘fuzzy’) region ^34^. This ‘defuzz’ parameter must be specified when combining - ve and +ve sense annotated events, since this ensures that the coordinates are reported in a consistent manner regardless of the directionality of the original RNAseq reads aligned to each recombination event.

#### Plot_CS_Freq.py

Once all the viral recombination events are reported in a single BED file using a standard coordinate system and ensuring the directionality of the RNAseq method is properly accounted for, the abundance of different groups of recombination events are reported using the ‘JFreq’ metric, as previously described ^21^. JFreq is calculated by taking the number of sequence reads mapping to a single event, or group of recombination events, and normalizing to the number of reads mapping to the whole virus genome in each dataset. The number of mapped reads is determined using the samtools ‘depth’ command, invoked automatically by ViReMa (v0.29 or greater) and which is stored in the ‘[root]_report.txt’ file. JFreq is output as the number of junctions detected per 10’000 mapped reads.

This final script outputs the JFreq values for the majors groups of recombination events, including: sgmRNAs (defined as any recombination event with a start site prior to nucleotide coordinate of 80); insertions (where the stop site is found upstream of the start site); deletions (where the stop site is found downstream of the start site); and micro-insertions or micro-deletions (where the size of the insertion or deletion is smaller than or equal to 5 nucleotides in length). Further, the JFreqs are broken down for each of the established sgmRNAs for SARS-CoV-2 if the acceptor site of the recombination events is found at their respective known junctions using the WA-1 coordinates for each sgmRNAs (Spike: 21557, ORF3a: 25386, E:26238, M:26474, ORF6:27042, ORF7a: 27389, ORF7b: 27761, ORF8: 27889, N: 28261). Any remaining sgmRNAs found at other sites are grouped and reported as ‘non-canonical’ sgmRNAs.

Each of these JFreq values are output to a final report file for each sample analysed using this batch script. As such, multiple individual samples can be cross compared to evaluate for changes in abundance of RNA recombination events as well as each specific type of recombination event.

### Phylogenetic Analysis

Phylogenetic tree analysis using UShER and visualization using Nextstrain was done under the CZ Gen EPI tool^72,79^. The CZ Gen EPI which is maintained by the Chan Zuckerberg Initiative and enabled by data from GenBank allows the generation and annotation of phylogenetic trees in Nextstrain. UShER provides a faster and more robust real-time analysis of the SARS-CoV-2 pandemic by utilizing genomes from GISAID, GenBank, COG-UK, and CNCB ^79^. We uploaded our multifasta files onto CZ Gen EPI tool and build trees through the UShER option. Once the trees were completed, they were visualized on Nextstrain, and annotations on the samples included^79^.

### Informed Consent Statement

Patient consent was waived due to the nature of the activity, which was a response to the pandemic.

### Data Availability Statement

Data The Kenyan SARS-CoV genome sequence data used had been submitted to either global initiative on sharing avian influenza data (GISAID, https://www.gisaid.org/ accessed on 10 January 2022 or (NCBI, https://www.ncbi.nlm.nih.gov/).

### Data Availability Statement

ViReMa bash script for recombinant RNA species with the current submission is available on GitHub https://github.com/andrewrouth/ARTIC_ViReMa/tree/main. Any updates will be published on GitHub and the final version will be cited in the manuscript.

## Author contributions

D. L.: Designed the study, analyzed data, performed data visualization, and first manuscript draft, B.M.: Analyzed the data and edited the manuscript, G.K. and K.M: Curated and analyzed the data, S.O., E.K., P.D., C.M. and R.N.: designed protocols, processed and sequenced the samples, T.D.O. and M.K.N: applied for funds, revised and edited the manuscript, A.R.: designed the analysis software, revised and edited the manuscript, S.O.O.: Conceptualized and designed the study, analyzed data, applied for funds, supervised the project, revised and edited the manuscript.

## Institutional Review Board Statement

The study was conducted in accordance with the Declaration of Helsinki, and approved ILRI Institutional Research Ethics Committee (ILRI-IREC2020-52), The COVID-19 surveillance and testing data were collected from public database of the Kenya Ministry of Health (KMOH) with administrative approval from the ministry.

## Acknowledgements

We wish to acknowledge the Kenya county surveillance teams and ILRI genomic team for supporting COVID-19 sample collection and processing.

