## Supplementary Figures for "Characterization of SARS-CoV-2 intrahost genetic evolution in vaccinated and non-vaccinated patients from the Kenyan population"

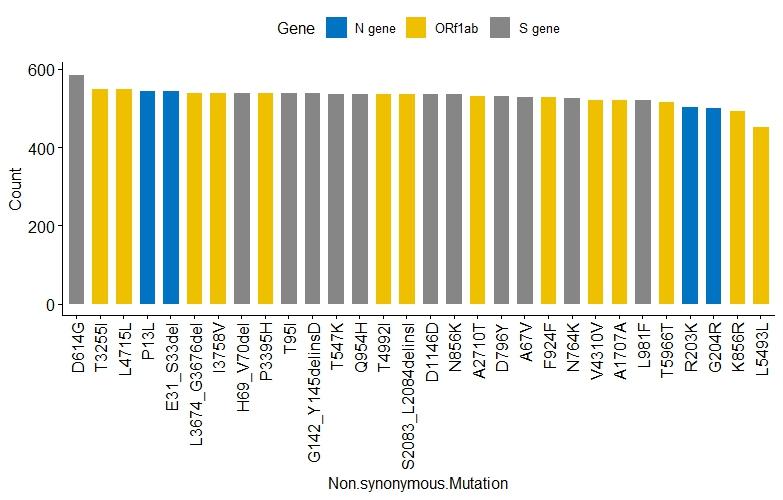

**Supplementary Fig. 1: Top 30 mutations in non-vaccinated and vaccinated patients in Kenya.**

**Counts**

**Mutations**

Supplementary Fig. 1: Shows the top 30 mutations in non-vaccinated and vaccinated patients in Kenya. Mutations were mostly found on the ORF1a/b, S, and N gene. Mutations in blue are found in the N genes, those in yellow are found in the ORF 1a/b, and those in grey are found in the S gene.

**Supplementary Fig. 2: Recombination events per patient in non-vaccinated and vaccinated patients in Kenya.**

***Deletion***

***Duplication***

***Insertions***

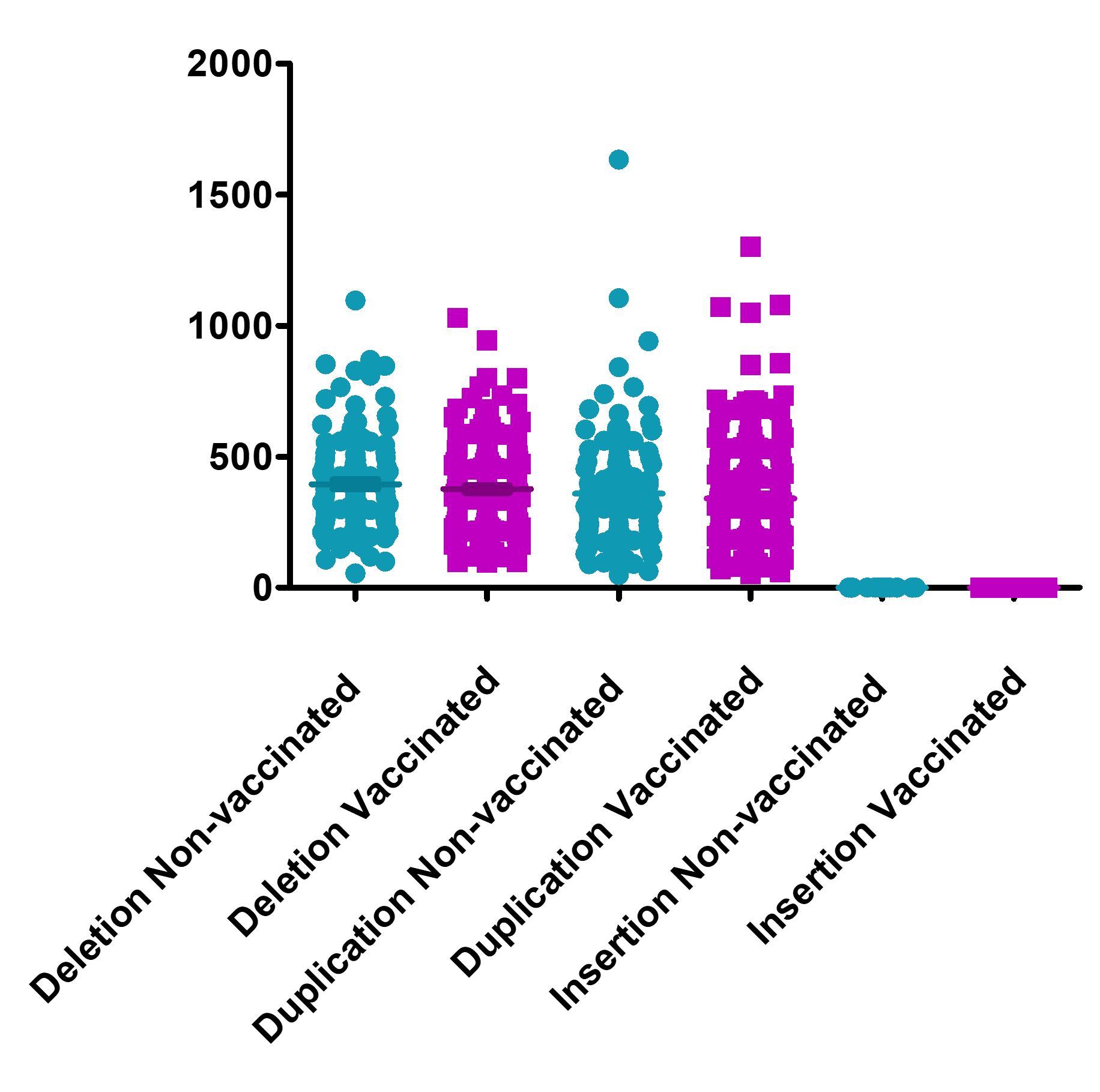

**Non-vaccinated**

**Vaccinated**

**Patient samples**

**Recombination events**

Supplementary Fig. 2: Shows the number of deletion, duplications, and insertion events per patient in non-vaccinated and vaccinated patients in Kenya. Green circles represent recombination events found in non-vaccinated patients and purple boxes represent those found in vaccinated patients.

**5’ UTR**

**ORF1a**

**ORF1b**

**S**

**ORF3a**

**ORF6**

**E**

**M**

**ORF7a**

**ORF8**

**N**

**ORF10**

**3’ UTR**

**log_10_ no. of recombination events**

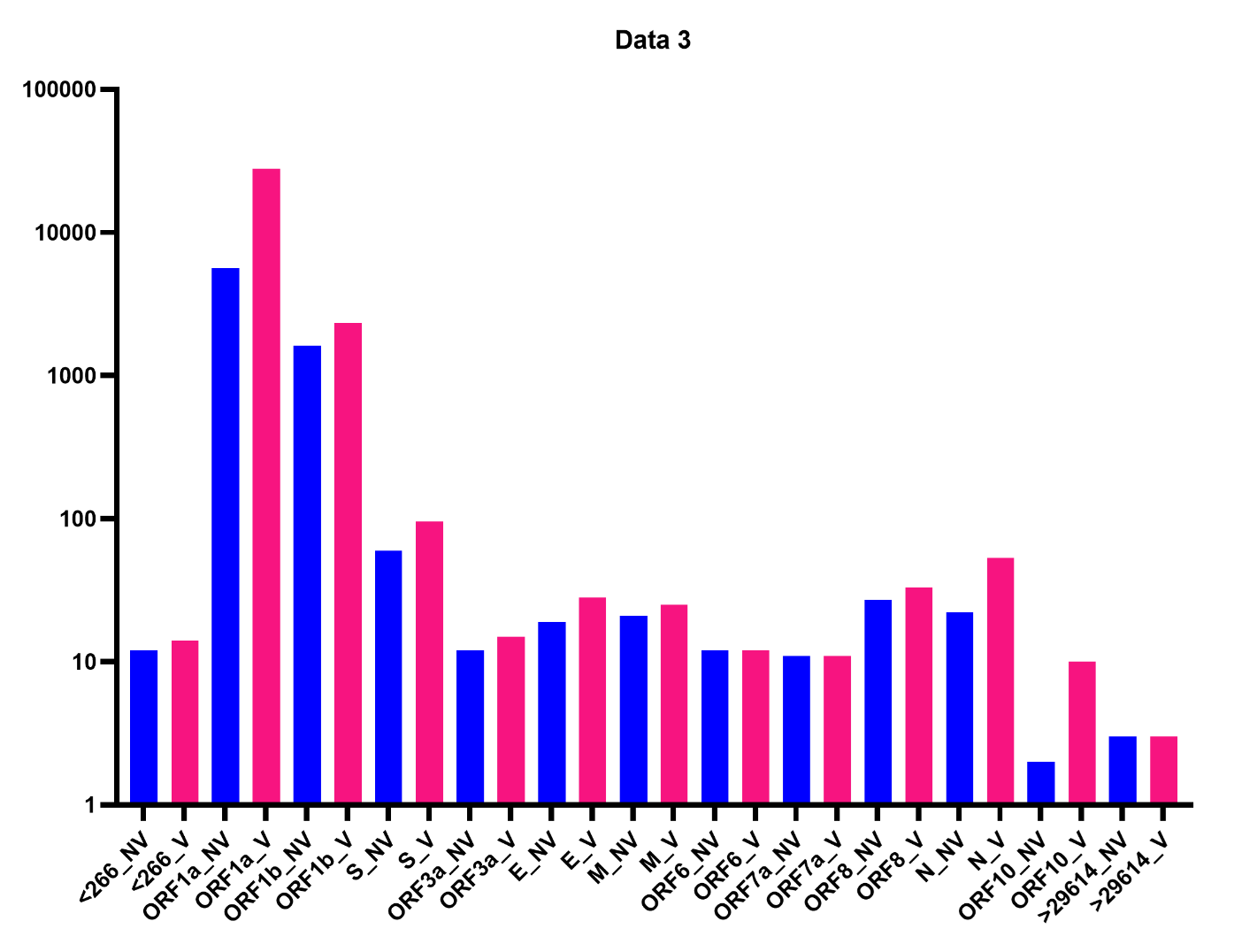

**Deletion regions**

**70 – 21545 (S gene)**

**70 – 26447 (M gene)**

**70 – 25358 (ORF3a)**

**71 – 27389 (ORF7a)**

**Non-vaccinated counts**

**20917**

**19597**

**3796**

**3194**

**402**

**398**

**353**

**673**

**570**

**10489**

**10057**

**2680**

**1742**

**369**

**323**

**277**

**265**

**251**

**12769 - 13315 ( DVG)**

**4640 - 9784 ( DVG)**

**7001 - 11654 ( DVG)**

**21737 - 21771 ( DVG)**

**70 – 28240 (N gene)**

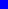

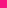

**Non-vaccinated**

**Vaccinated**

**Vaccinated counts**

**A.**

**B.**

**Supplementary Fig. 3: Top recombination events between non-vaccinated and vaccinated individuals.**

**C.**

Supplementary Fig. 3: Top recombination events between non-vaccinated and vaccinated individuals. A. Top recombinant RNA species between vaccinated and non-vaccinated individuals and the counts. B & C. Recombination events based on major genome positions between vaccinated and non-vaccinated individuals.

Supplementary Fig. 4: The boxplots represent the JFreq (junction frequency) quantification of the recombination RNA species between sex (male and female), in the cohort of vaccinated and non-vaccinated individuals. Statistical significance was determined using one way ANOVA and Tukey multiple comparison tests.

**Supplementary Fig. 4: JFreq (junction frequency) quantification of recombinant RNA species between sex (male and female), in the cohort of vaccinated and non-vaccinated individuals.**

***ns***

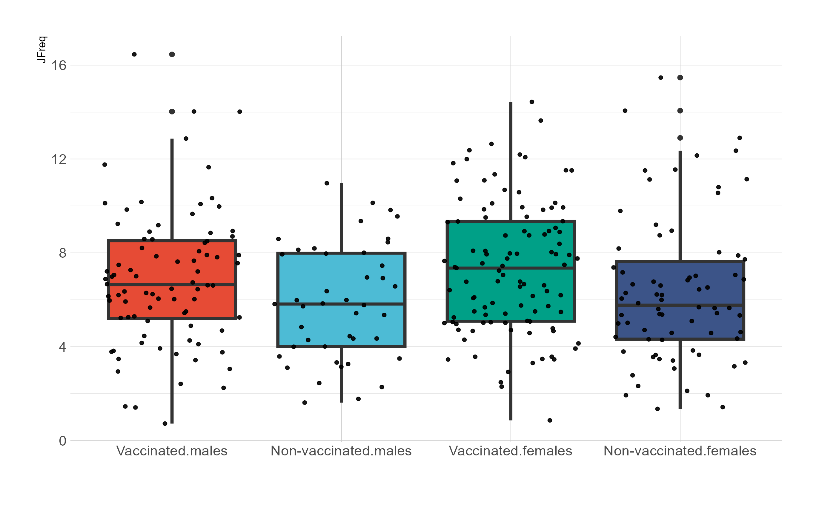

**Non-vaccinated**

**females**

**Vaccinated**

**females**

**Vaccinated**

**males**

**Non-vaccinated**

**males**

**0**

**4**

**8**

**12**

**16**

**JFreq**

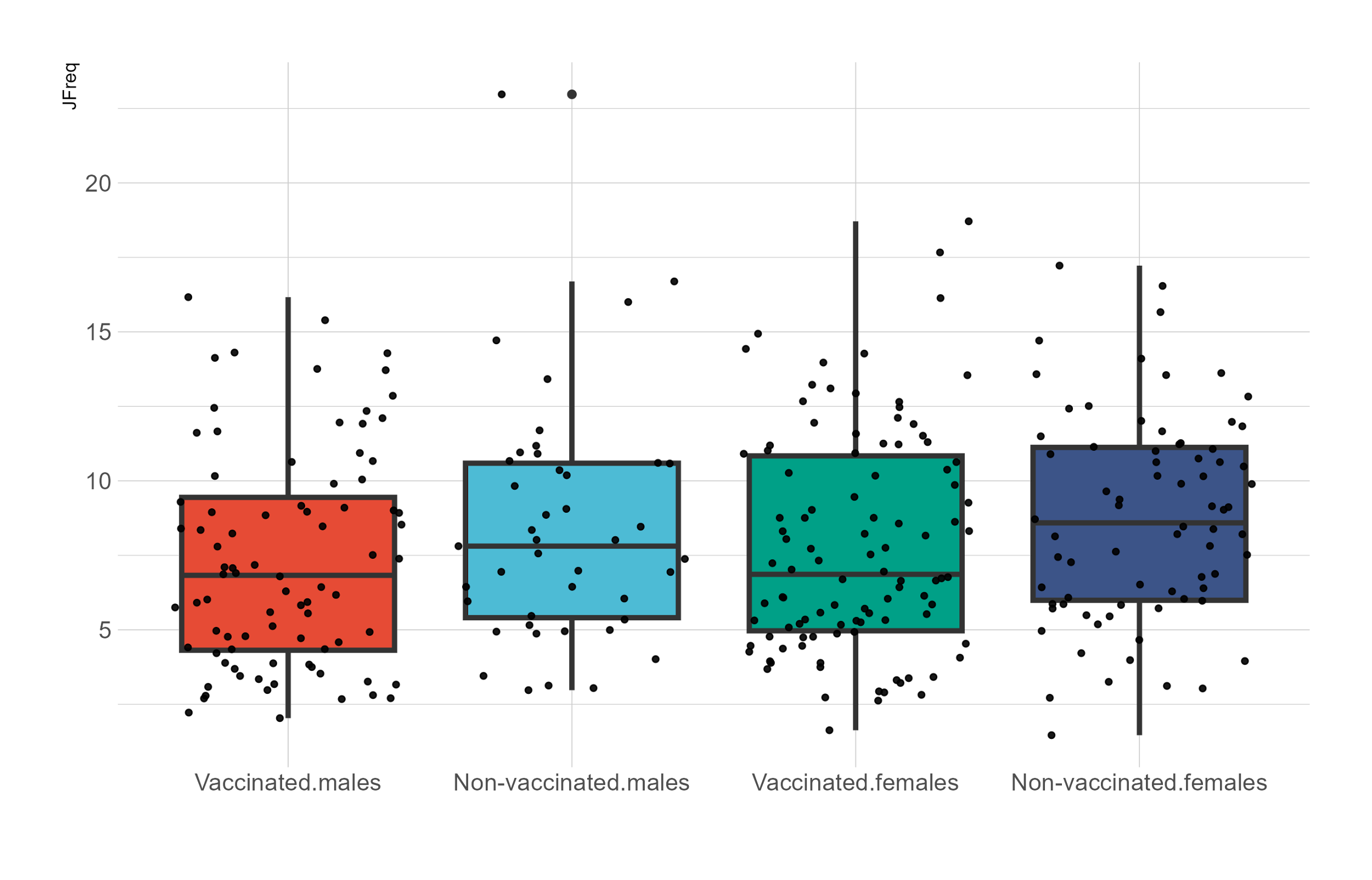

**5**

**10**

**15**

**20**

**Vaccinated**

**males**

**Non-vaccinated**

**males**

**Vaccinated**

**females**

**Non-vaccinated**

**females**

**JFreq**

**Sub-genomic RNAs**

**Large insertions**

**Micro-deletions**

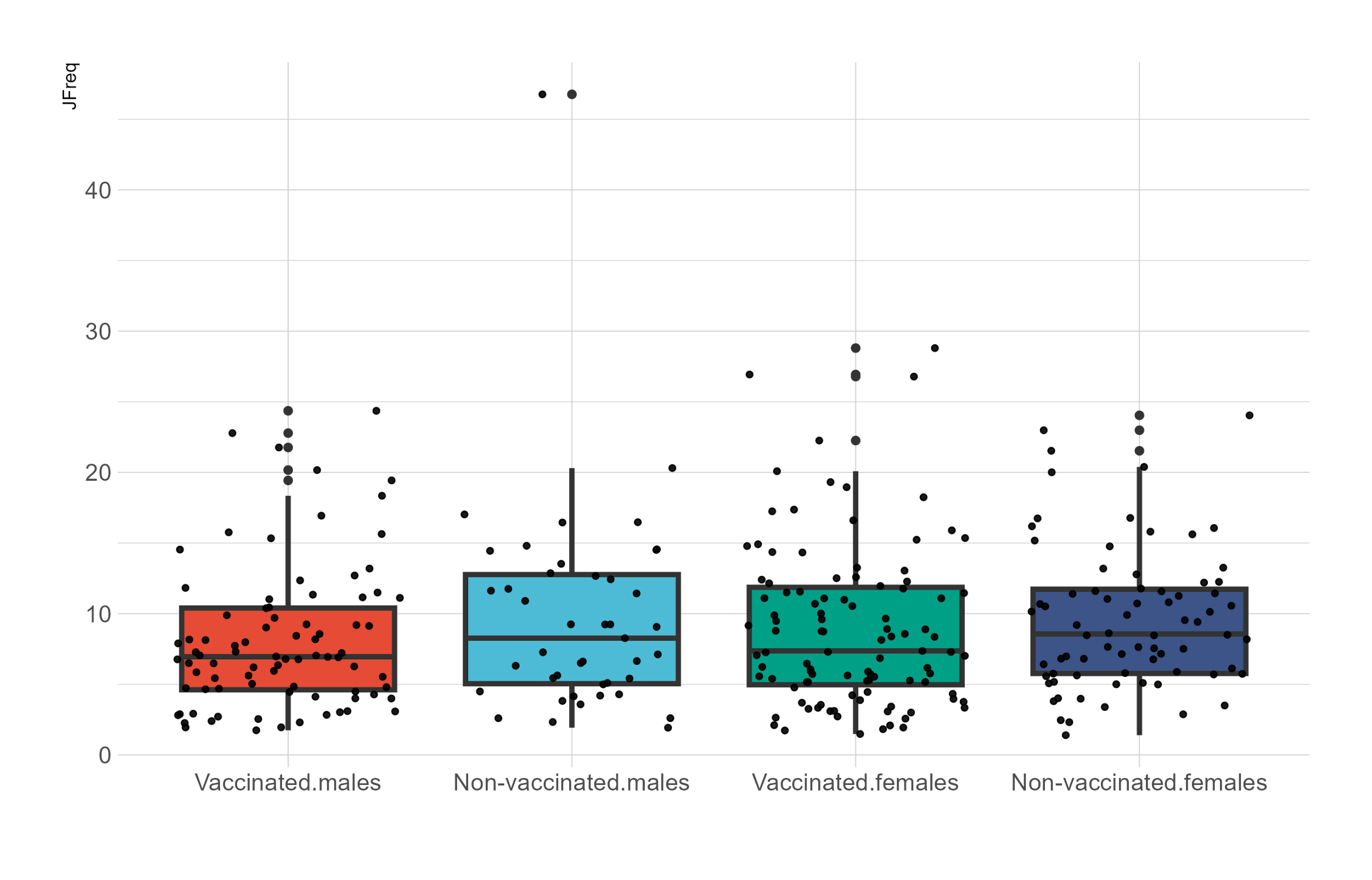

**0**

**10**

**20**

**30**

**40**

**Vaccinated**

**males**

**Non-vaccinated**

**males**

**Vaccinated**

**females**

**Non-vaccinated**

**females**

**JFreq**

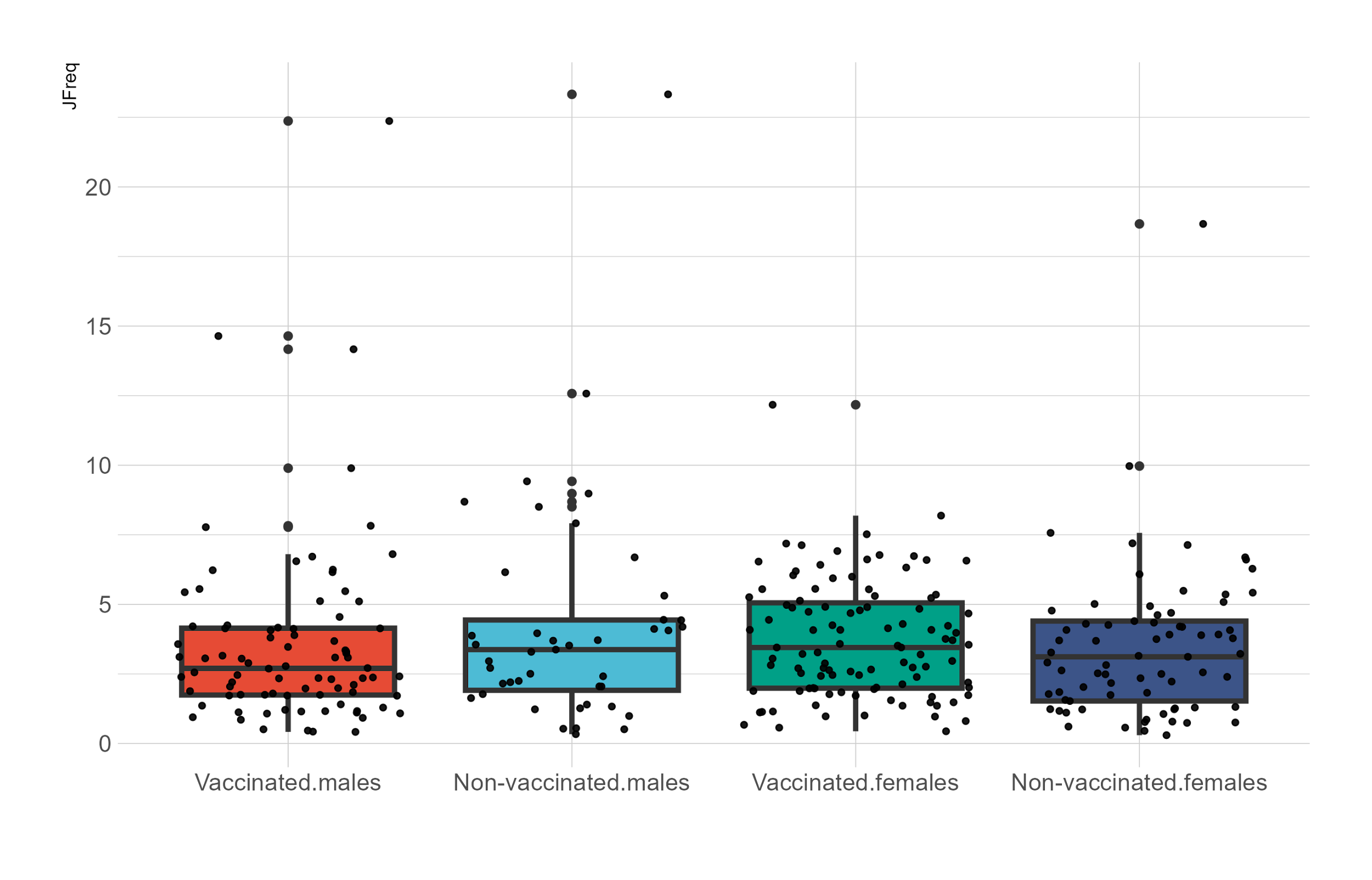

**0**

**5**

**10**

**15**

**20**

**Vaccinated**

**males**

**Non-vaccinated**

**males**

**Vaccinated**

**females**

**Non-vaccinated**

**females**

**JFreq**

**Defective RNA genomes**

**Micro-insertions**

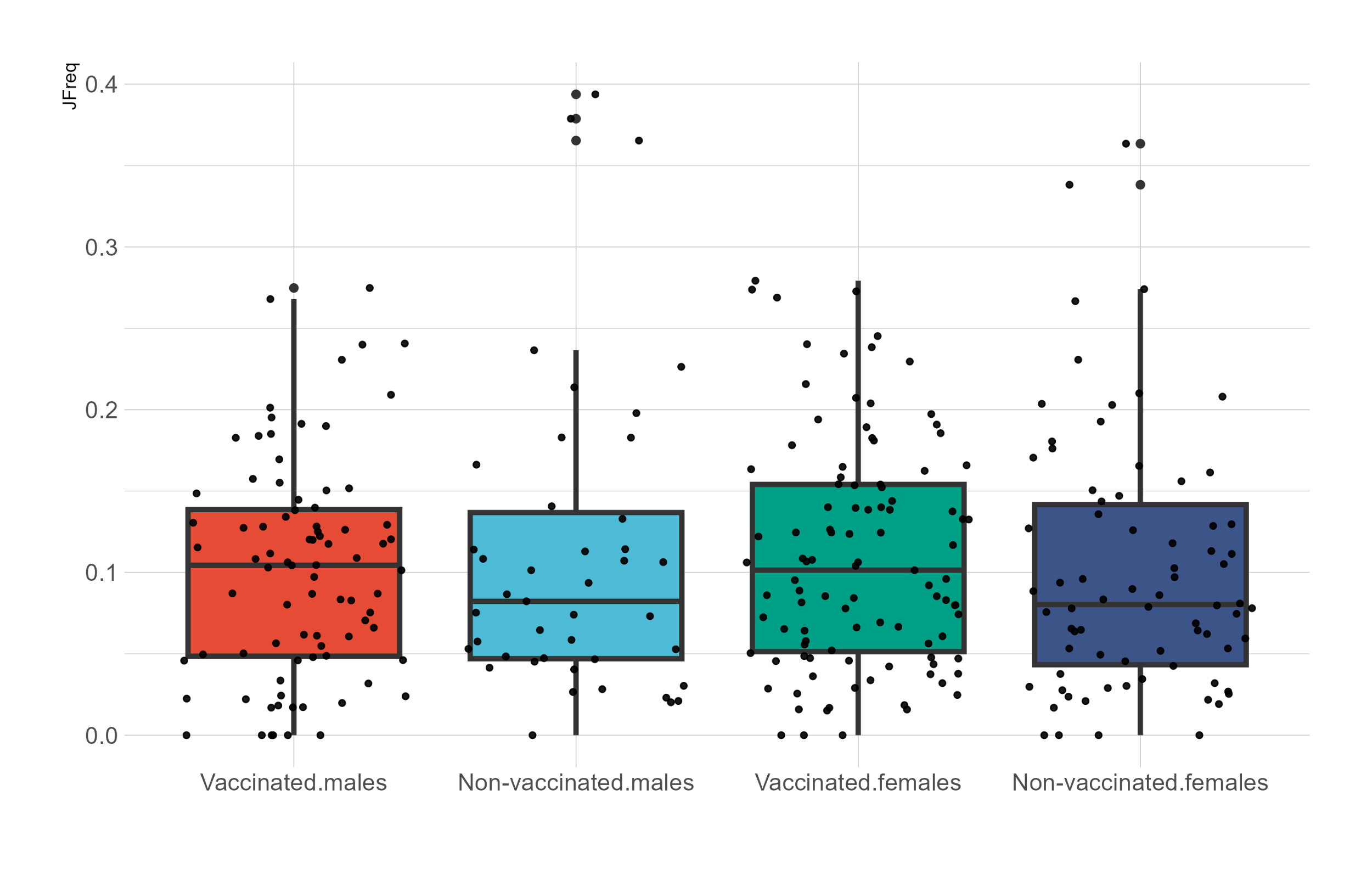

**Vaccinated**

**males**

**Non-vaccinated**

**males**

**Vaccinated**

**females**

**Non-vaccinated**

**females**

**0.0**

**0.4**

**0.3**

**0.2**

**0.1**

**JFreq**

***(F =3.345), p-value < 0.0195***

***ns***

***ns***

***ns***

**0**

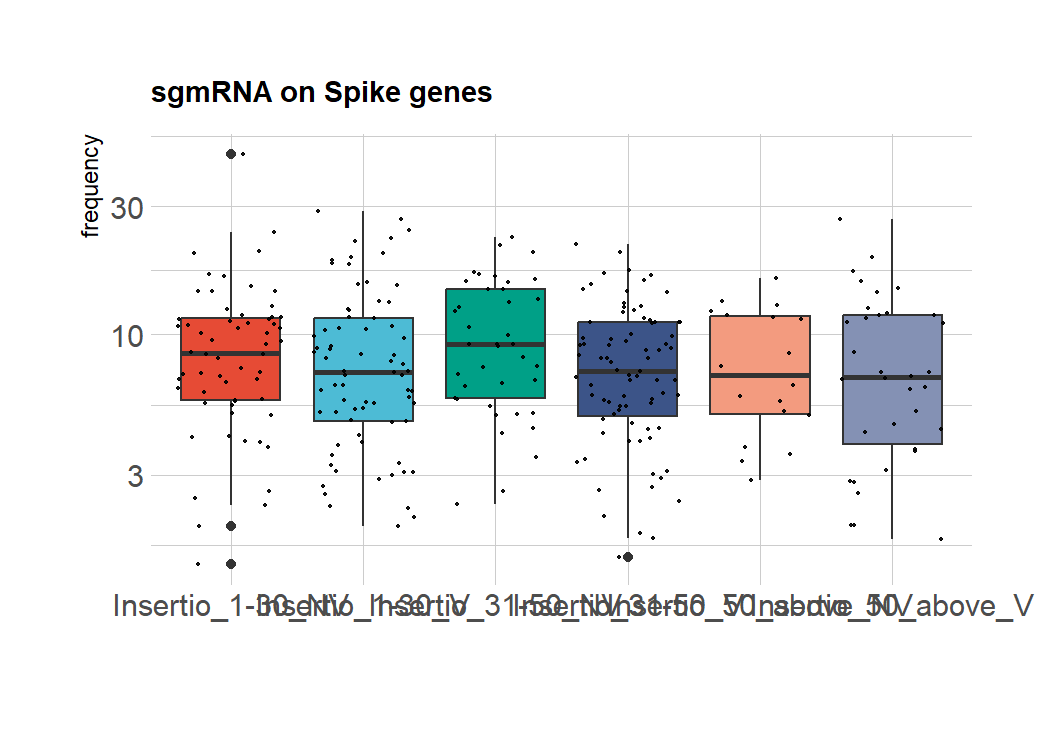

**JFreq**

**Sub-genomic RNAs**

**JFreq**

**Defective RNA genomes**

**Micro-insertions**

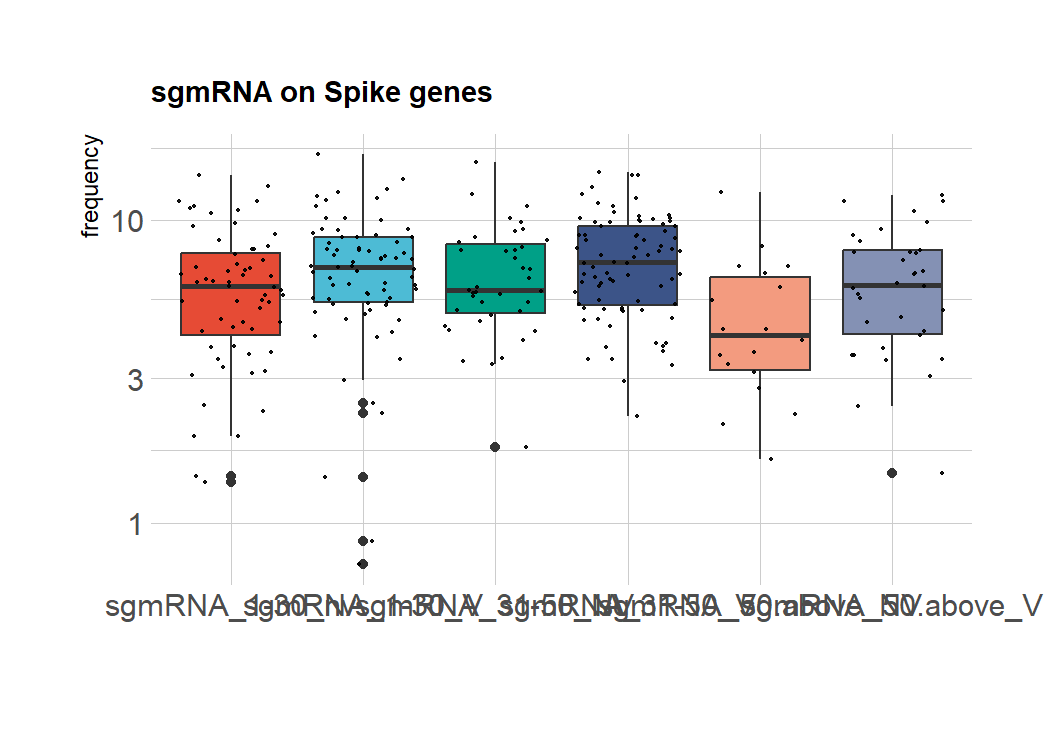

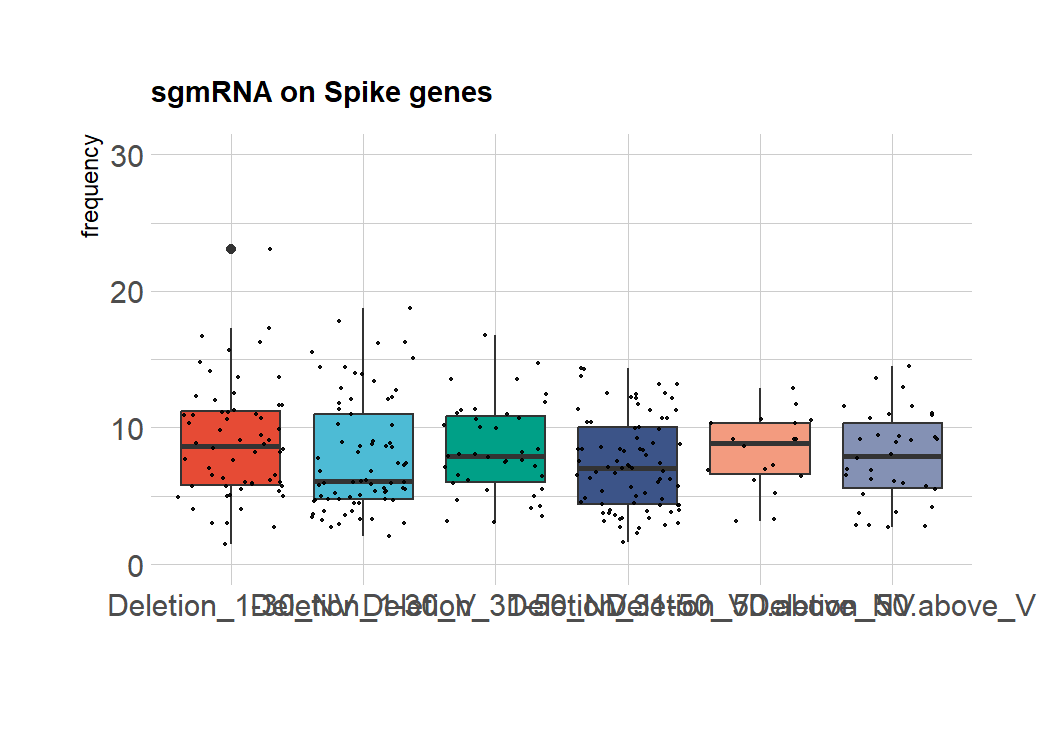

**Large insertions**

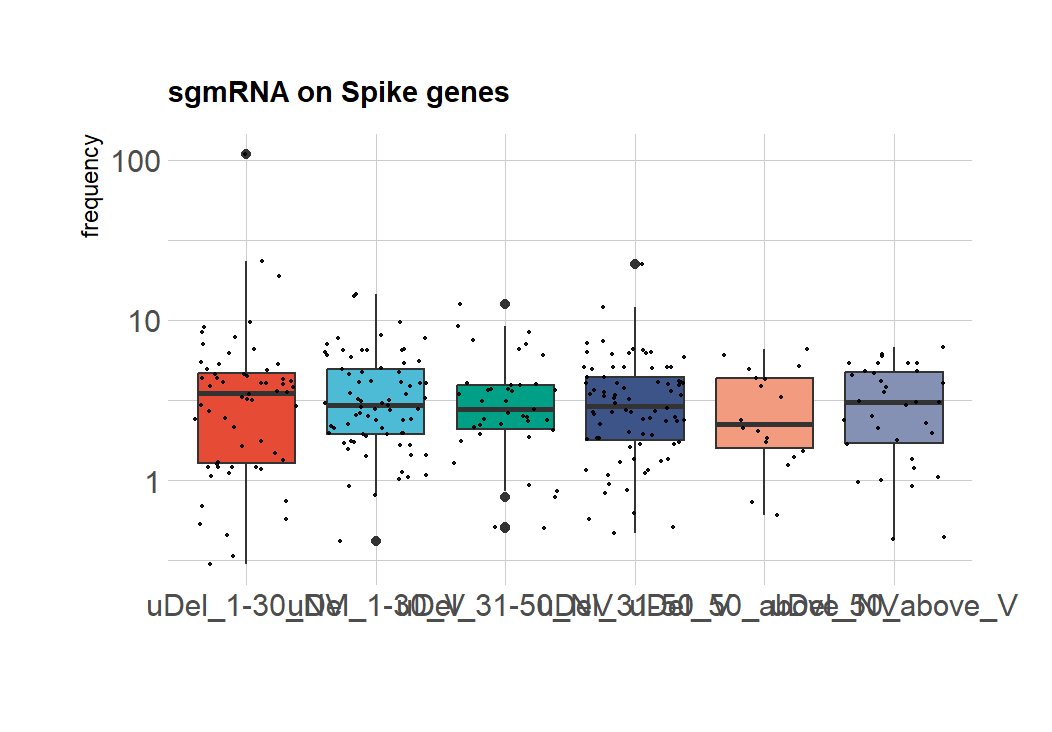

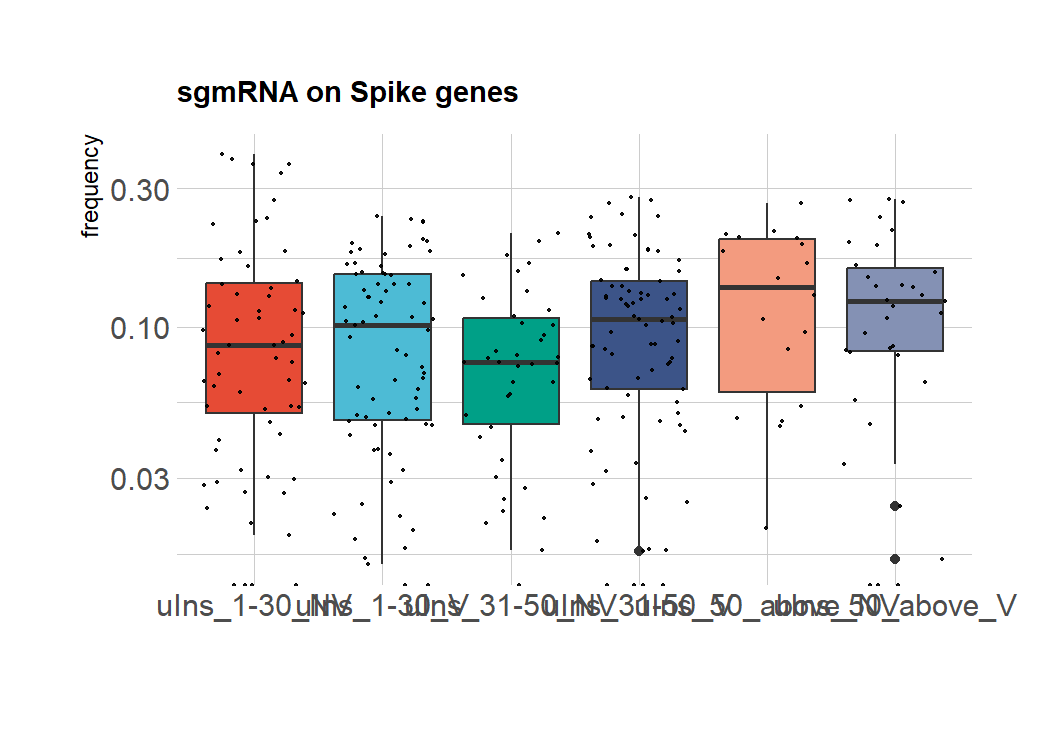

**JFreq**

**JFreq**

**JFreq**

**1-30**

**Non-vaccinated**

**1-30**

**Vaccinated**

**31-50**

**Vaccinated**

**>50**

**Vaccinated**

**31-50**

**Non-vaccinated**

**>50**

**Non-vaccinated**

**1-30**

**Non-vaccinated**

**1-30**

**Vaccinated**

**31-50**

**Vaccinated**

**>50**

**Vaccinated**

**31-50**

**Non-vaccinated**

**>50**

**Non-vaccinated**

**1-30**

**Non-vaccinated**

**1-30**

**Vaccinated**

**31-50**

**Vaccinated**

**>50**

**Vaccinated**

**31-50**

**Non-vaccinated**

**>50**

**Non-vaccinated**

**1-30**

**Non-vaccinated**

**1-30**

**Vaccinated**

**31-50**

**Vaccinated**

**>50**

**Vaccinated**

**31-50**

**Non-vaccinated**

**>50**

**Non-vaccinated**

**1-30**

**Non-vaccinated**

**1-30**

**Vaccinated**

**31-50**

**Vaccinated**

**>50**

**Vaccinated**

**31-50**

**Non-vaccinated**

**>50**

**Non-vaccinated**

**Micro-insertions**

*******

***p-value 0.05***

*******

***p-value 0.044***

***(F =3.366), p-value < 0.0057***

***ns***

***ns***

***ns***

***ns***

**Supplementary Fig. 5: JFreq (junction frequency) quantification of the recombination RNA species between age groups (1-30, 31-50, above 50), in the cohort of vaccinated and non-vaccinated individuals.**

Supplementary Fig. 5: The boxplots represent the JFreq (junction frequency) quantification of the recombination RNA species between age groups (1-30, 31-50, above 50) in the cohort of vaccinated and non-vaccinated individuals. Statistical significance was determined using one way ANOVA and Tukey multiple comparison tests.

**Supplementary Fig. 6: JFreq (junction frequency) quantification of sgmRNAs between complete, not complete vaccine dosage, in the cohort of vaccinated individuals.**

**.**

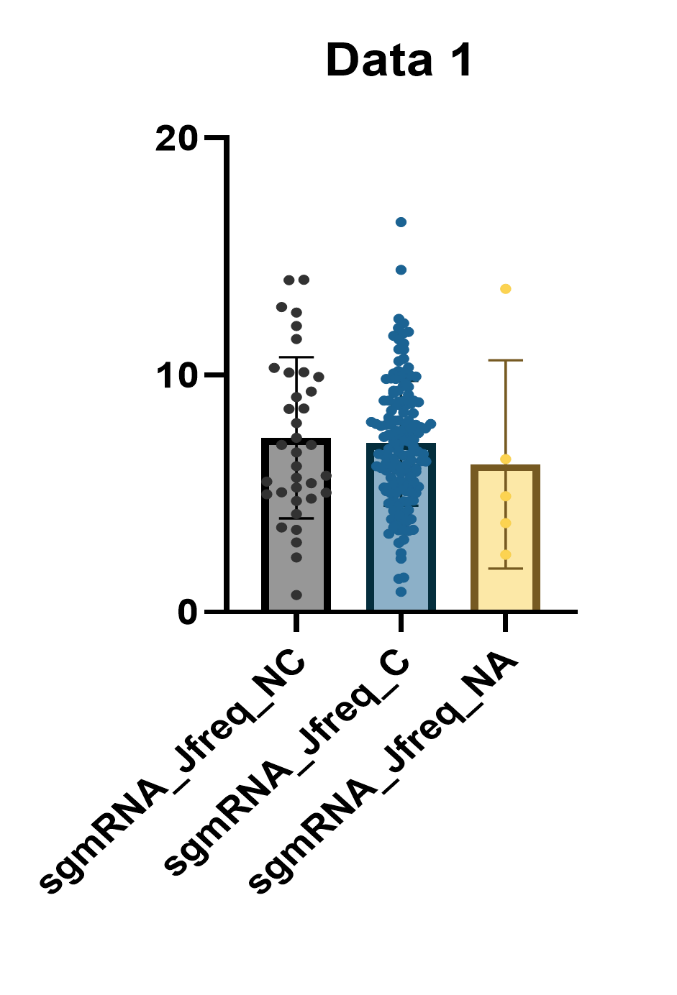

**Sub-genomic RNAs**

**Defective viral genomes**

**Large insertions**

**Micro-deletions**

**Micro-insertions**

**Not-Complete**

**Complete**

**NA**

**JFreq**

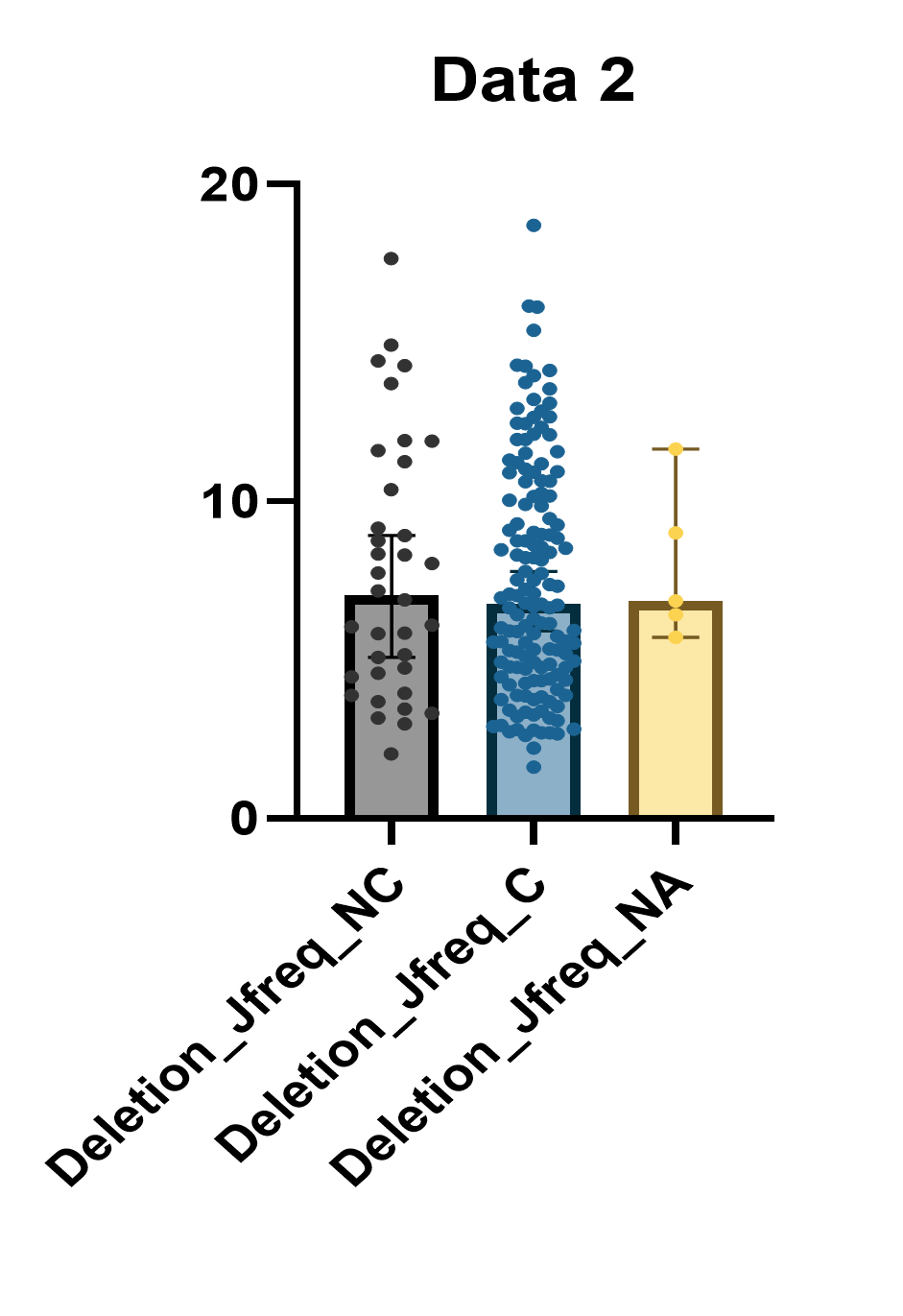

**Not-Complete**

**Complete**

**NA**

**JFreq**

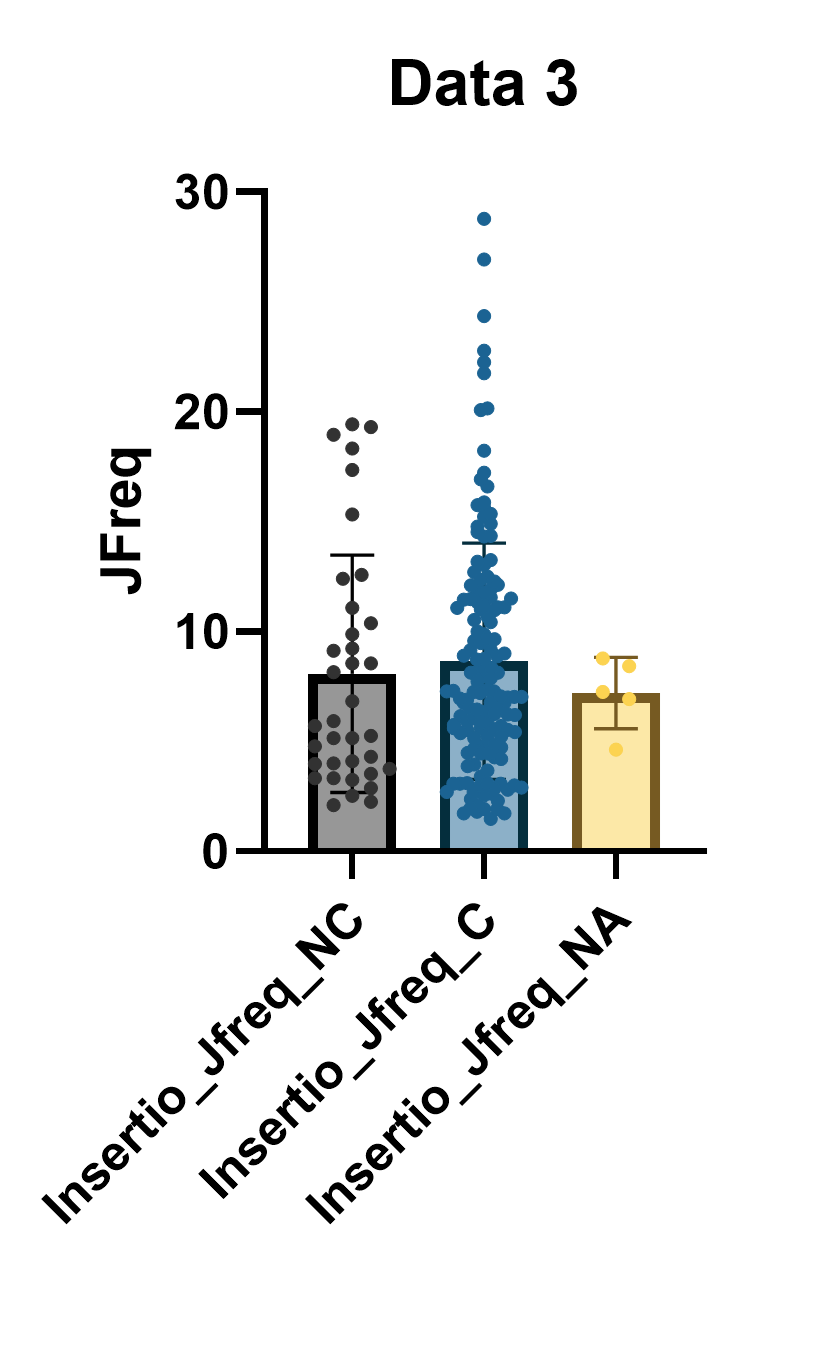

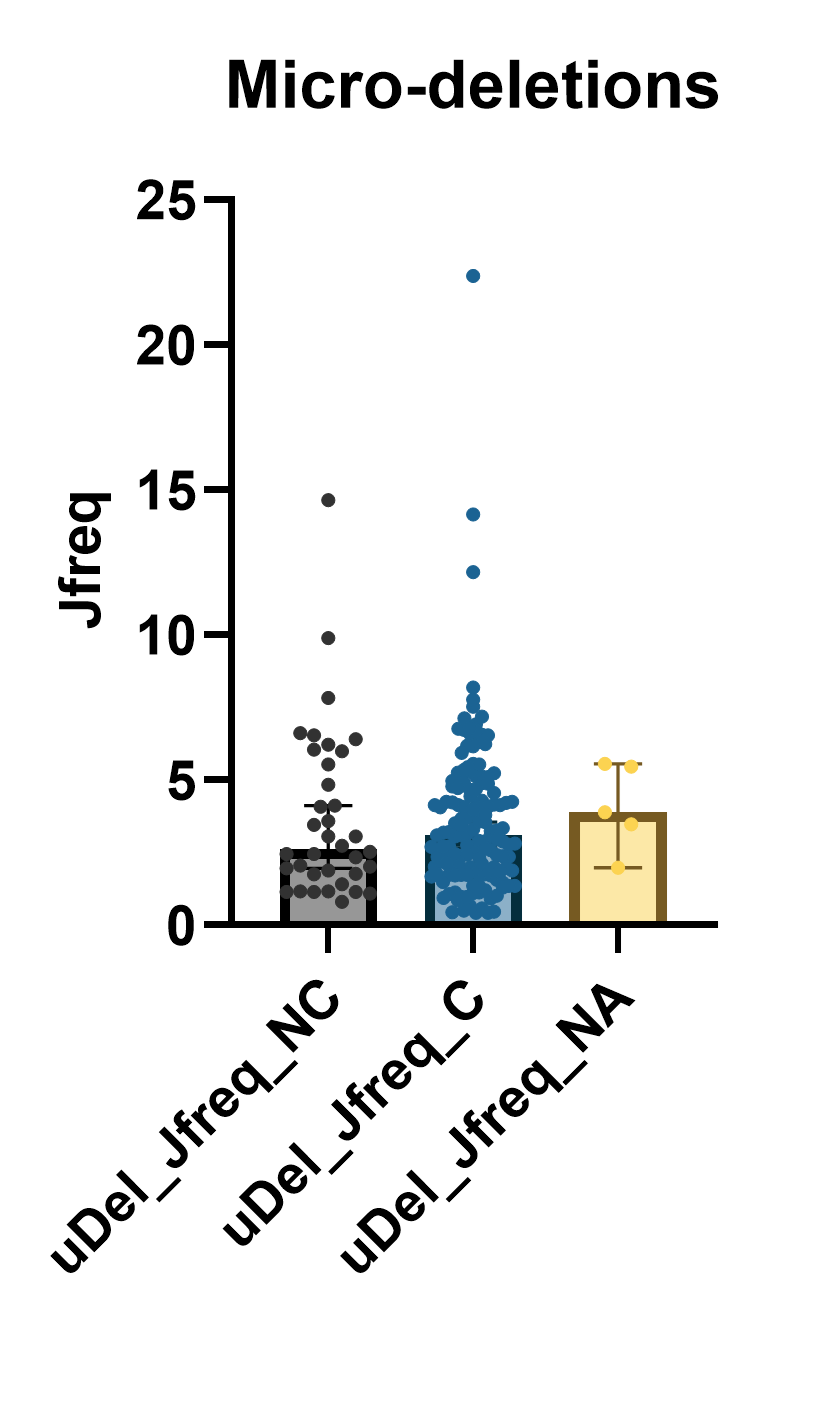

**Not-Complete**

**Complete**

**NA**

**Not-Complete**

**Complete**

**NA**

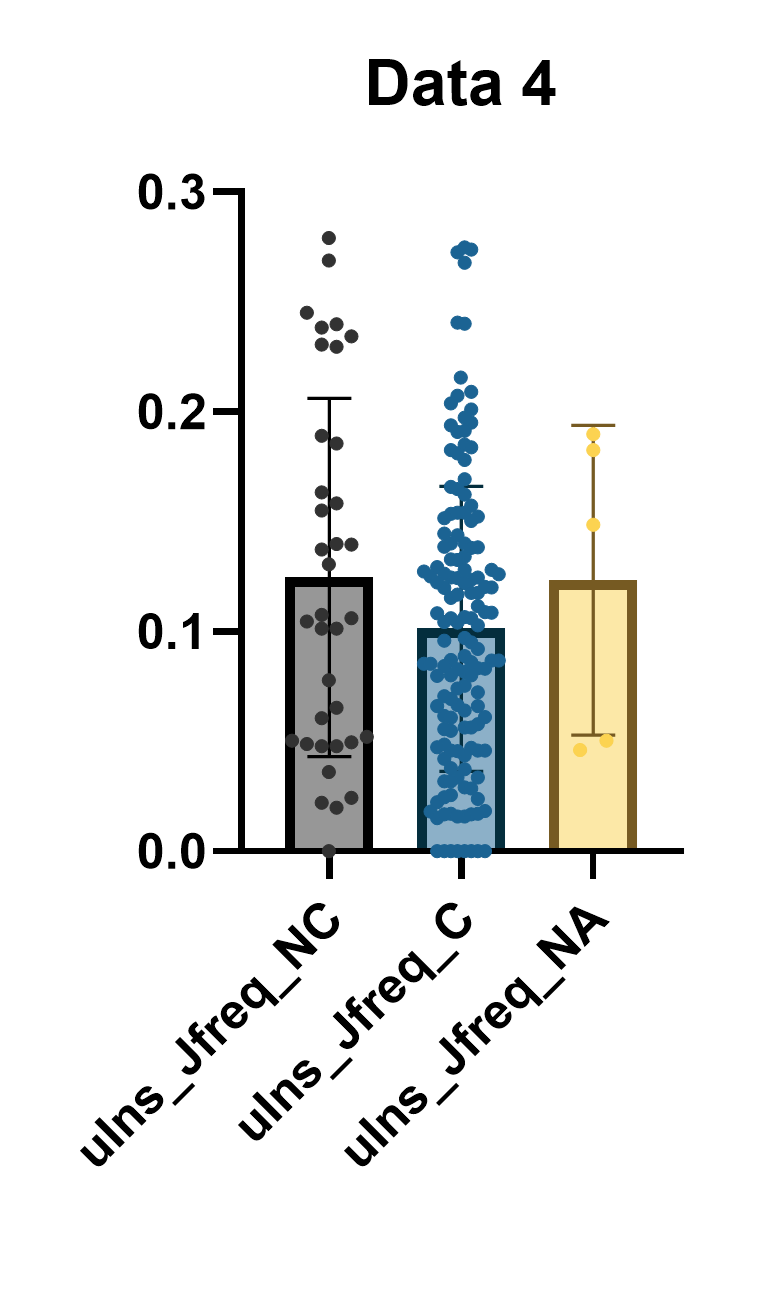

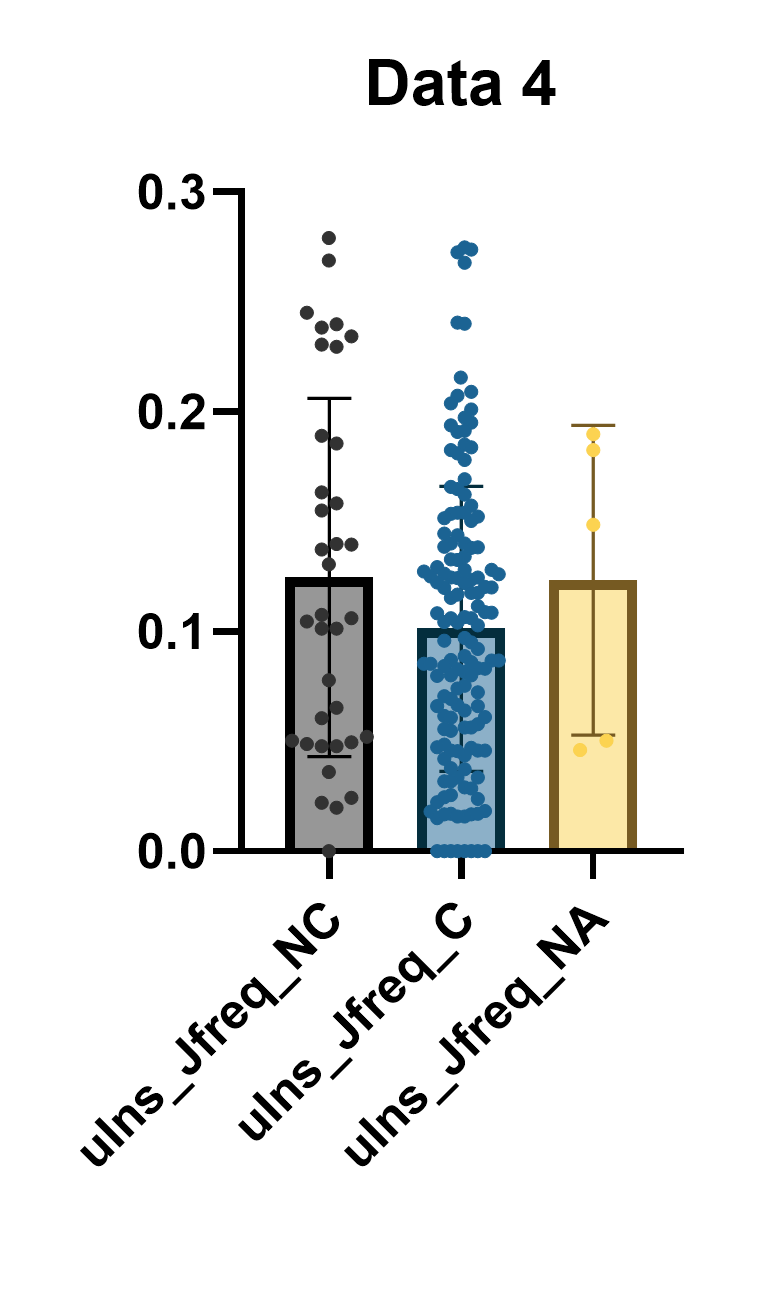

**JFreq**

**Not-Complete**

**Complete**

**NA**

**Dosage**

**Dosage**

**Dosage**

**Dosage**

**Dosage**

Supplementary Fig. 7: The boxplots represent the JFreq (junction frequency) quantification of the types of sgmRNAs between different sexes (male and female), in the cohort of vaccinated and non-vaccinated individuals.

**Spike**

**ORF3a**

**E**

**M**

**ORF7a**

**ORF8**

**Non-canonical**

**N**

**ORF7b**

**ORF6**

**Non-vaccinated females**

**Vaccinated**

**males**

**Non-vaccinated**

**Vaccinated females**

**Jfreq (log10)**

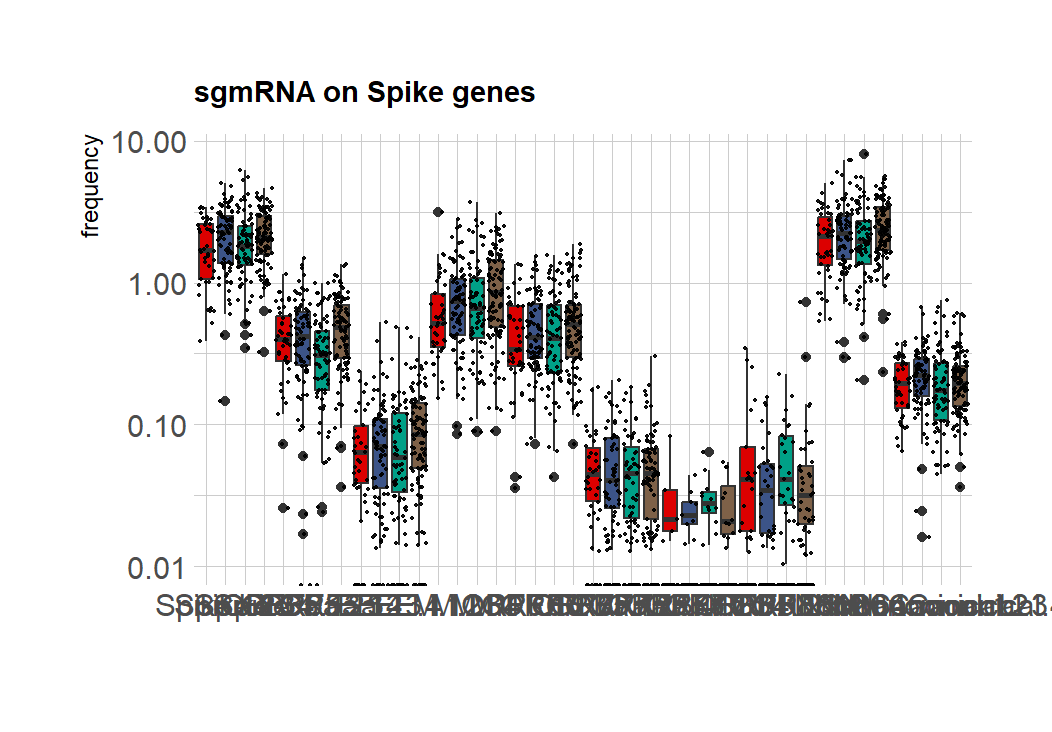

**Non-canonical**

**Supplementary Fig.7: JFreq (junction frequency) quantification of sgmRNAs between sex (male and female), in the cohort of vaccinated and non-vaccinated individuals.**

Supplementary Fig. 6: The boxplots represent the JFreq (junction frequency) quantification of the types of sgmRNAs between vaccine dosage , in the cohort of vaccinated individuals.

**Spike**

**ORF3a**

**E**

**M**

**ORF7a**

**ORF8**

**Non-canonical**

**N**

**ORF7b**

**ORF6**

**Spike**

**ORF3a**

**E**

**M**

**ORF7a**

**ORF8**

**Non-canonical**

**N**

**ORF7b**

**ORF6**

**Spike**

**ORF3a**

**E**

**M**

**ORF7a**

**ORF8**

**Non-canonical**

**N**

**ORF7b**

**ORF6**

**Non-vaccinated**

**Vaccinated**

**Age group 1-30**

**Age group above 50**

**Age group 31-50**

**JFreq(log10)**

**JFreq(log10)**

**JFreq(log10)**

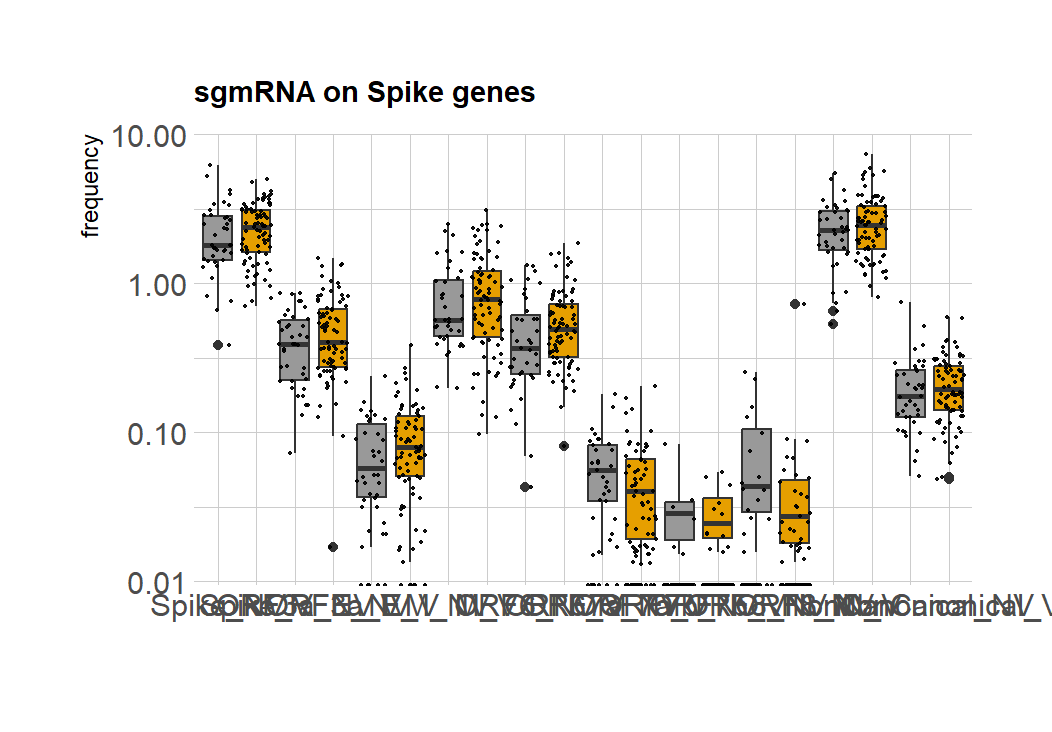

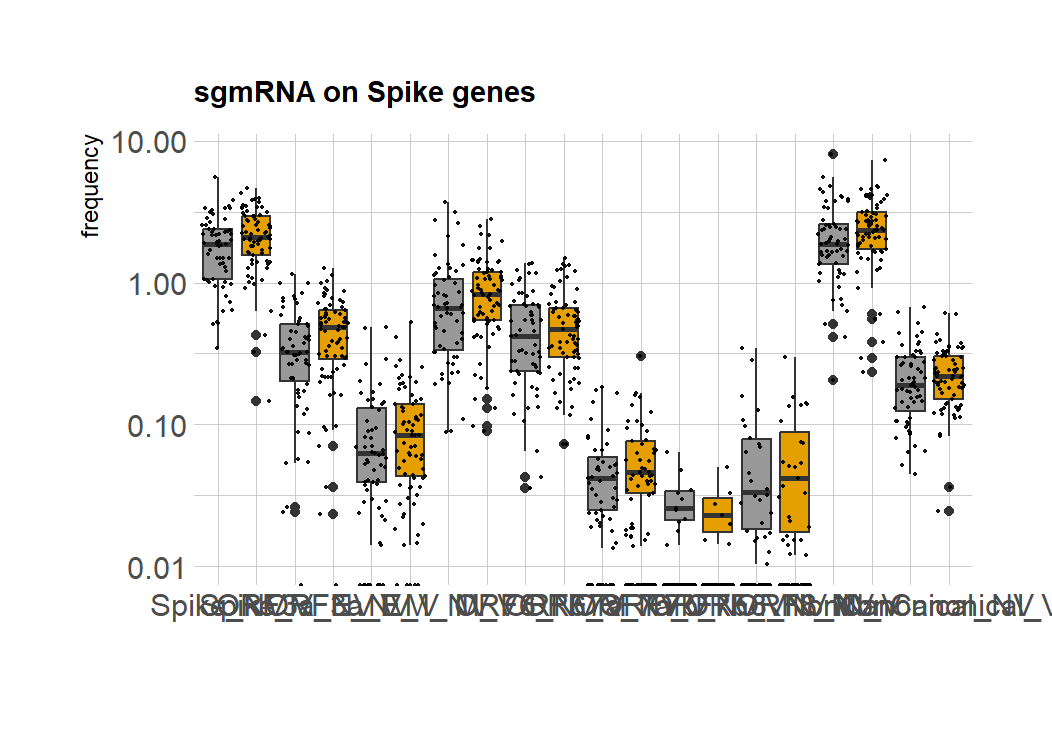

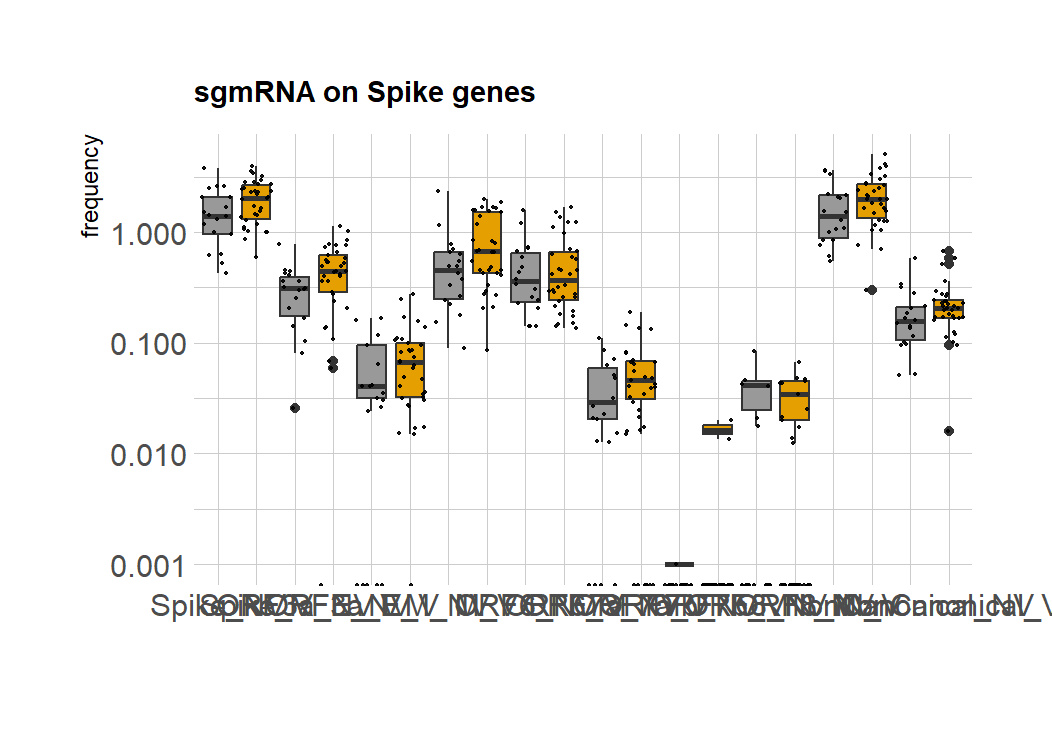

**Age group above 31-50**

**Supplementary Fig. 8: JFreq (junction frequency) quantification of sgmRNAs between age groups (1-30, 31-50, above 50), in the cohort of vaccinated and non-vaccinated individuals.**

Supplementary Fig. 8: The boxplots represent the JFreq (junction frequency) quantification of the types of sgmRNAs between age groups (1-30, 31-50, above 50) , in the cohort of vaccinated and non-vaccinated individuals.

**Supplementary Fig. 9: Box plot of genome coverage between waves and interwaves of SARS-CoV-2 from sequences in a Kenyan population.**

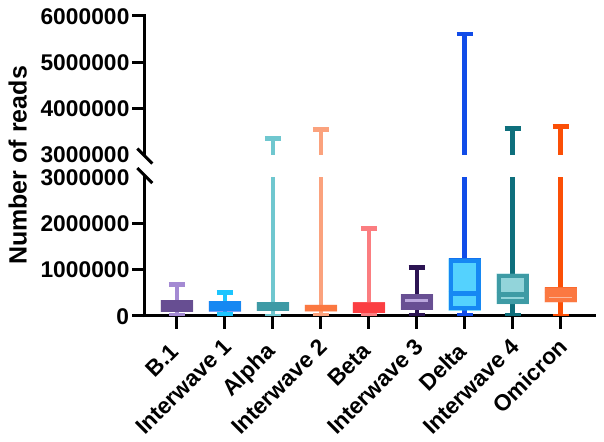

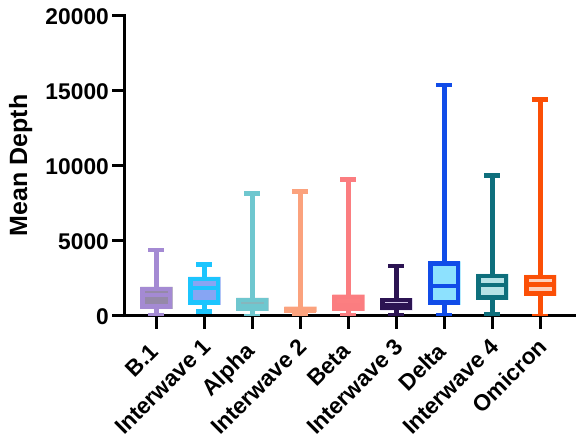

Supplementary Fig. 9: The boxplots represent the frequency of the number of reads and mean depth between all waves and interwaves.

**Supplementary Fig. 10 : ViReMa identifies recombination events between and during the peak of SARS-CoV-2 transmission waves.**

**B.1 , *n=78***

**IW1, *n=27***

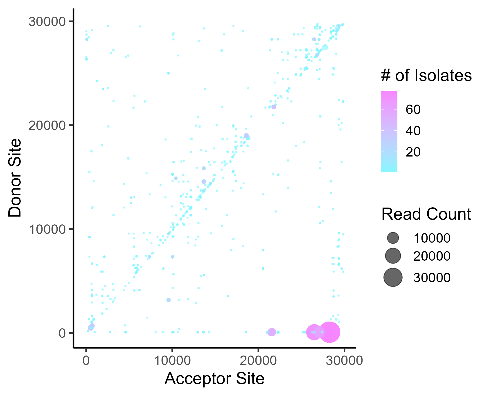

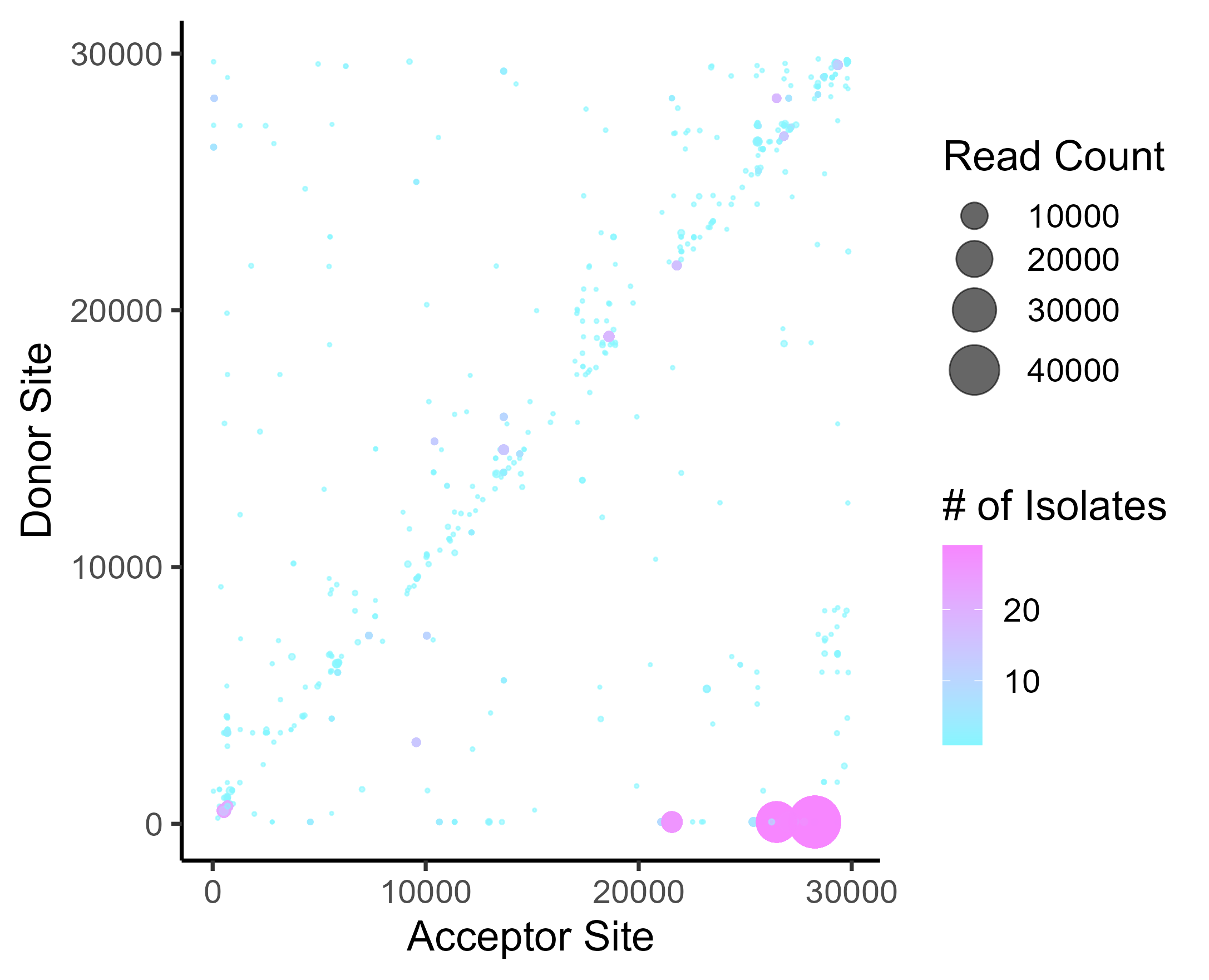

**Beta, *n=203***

**IW2 , *n=78***

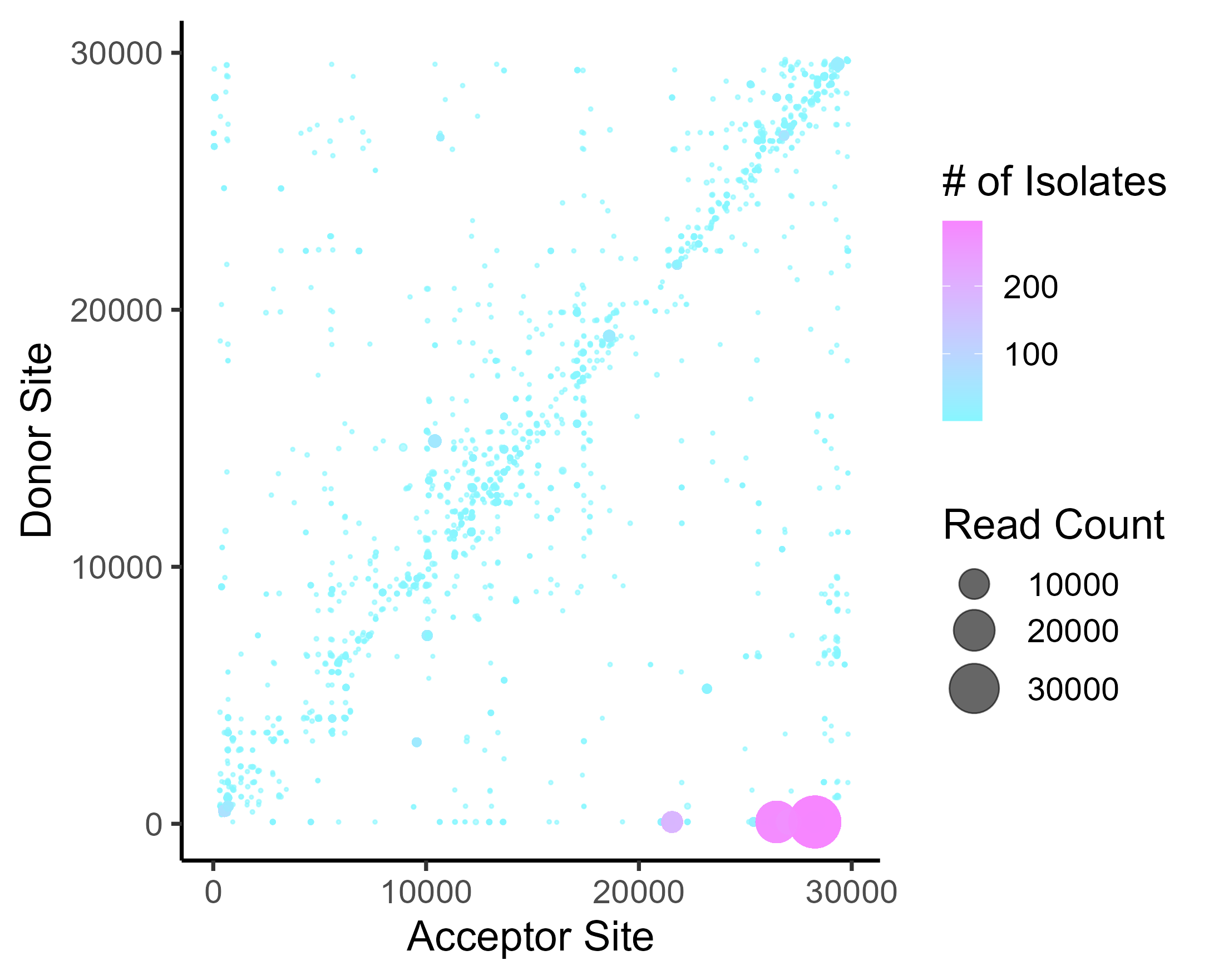

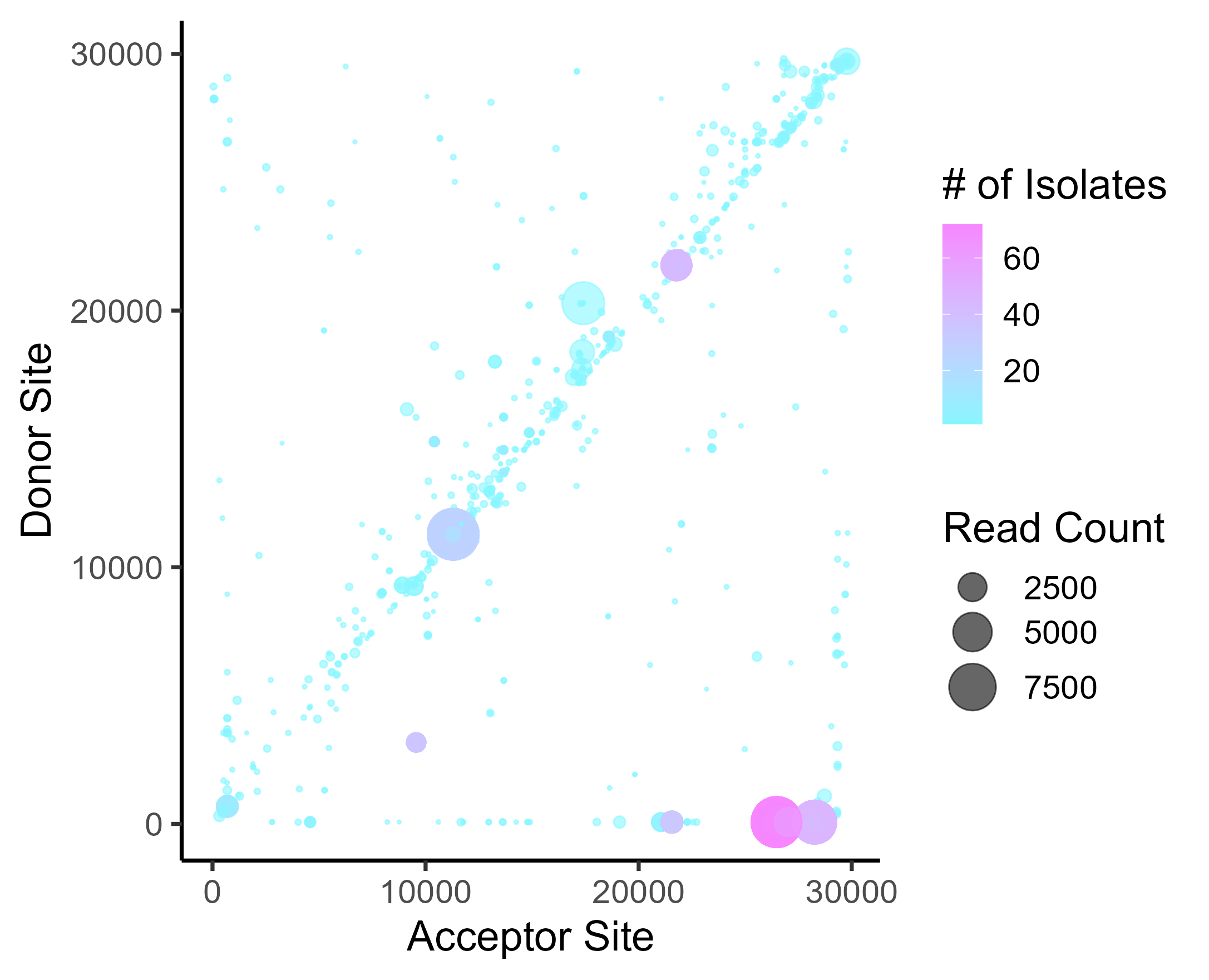

**Alpha, *n= 95***

**IW3, *n=35***

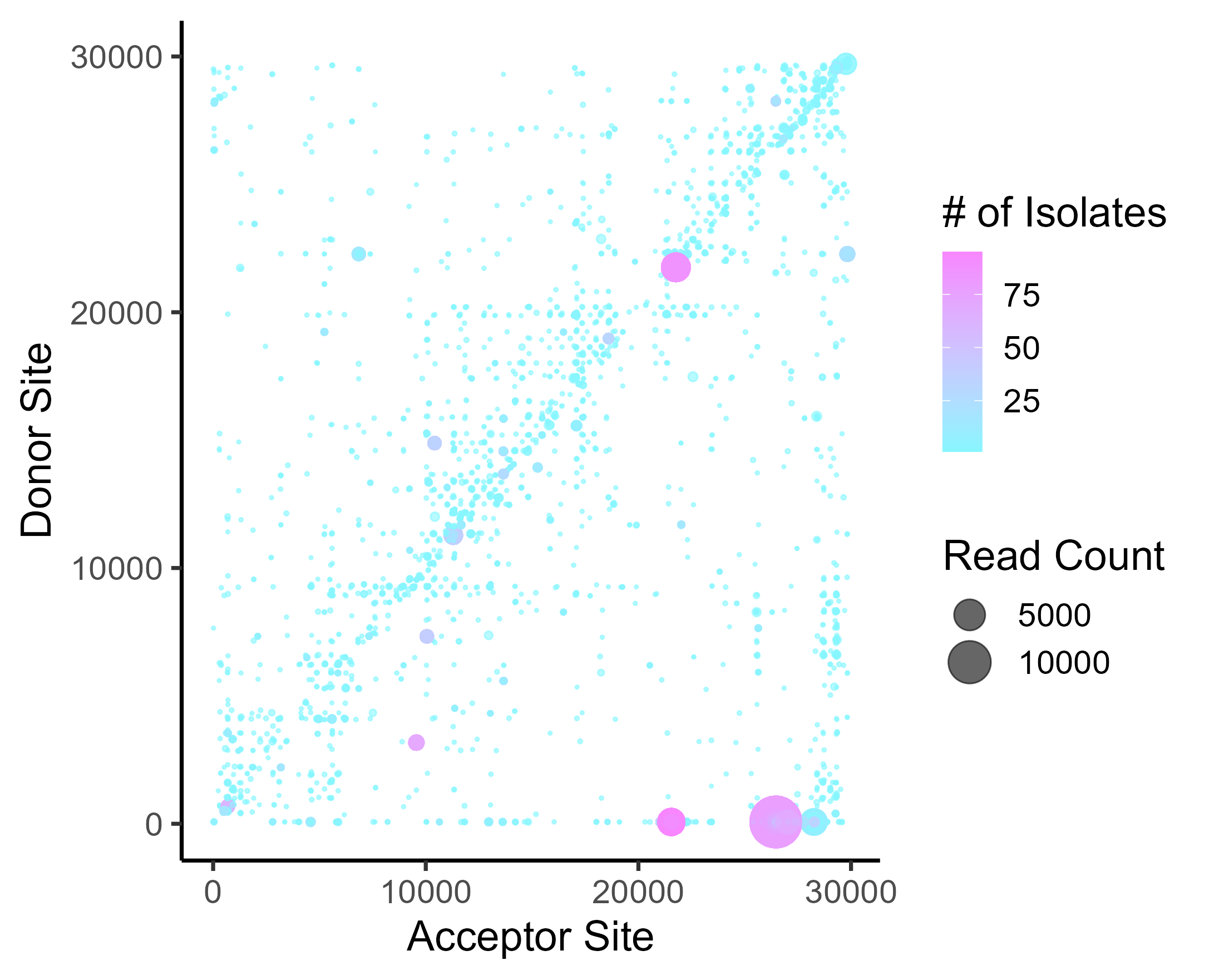

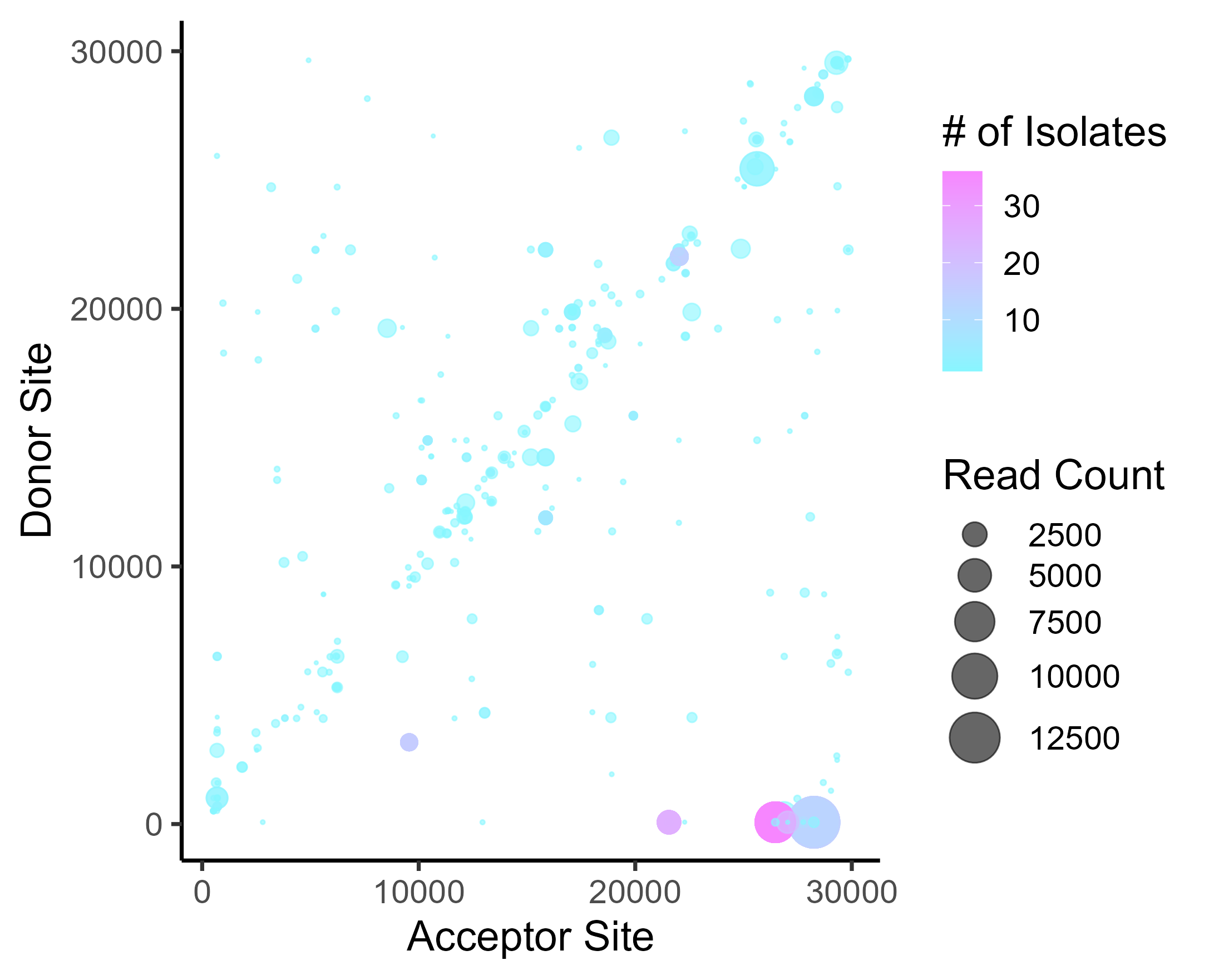

Supplementary Fig. 10: ViReMa scatter plots of SARS-CoV-2 recombination events and hotspots over B.1, interwave 1, Beta, interwave 2, Alpha, and interwave 3. The gradient in the scatter plot legend represents the number of patient samples containing a recombination event. The darker shaded circles in the scatter plot represent events that occur in multiple patient samples, while the circle size corresponds to the count of the reads of a recombination event.

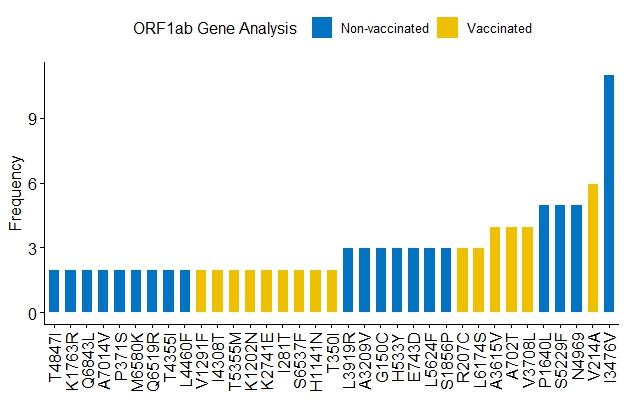

**Supplementary Fig. 11: Unique mutations in the ORF 1a/b, N, and S gene of non-vaccinated and vaccinated patients in Kenya.**

**A.**

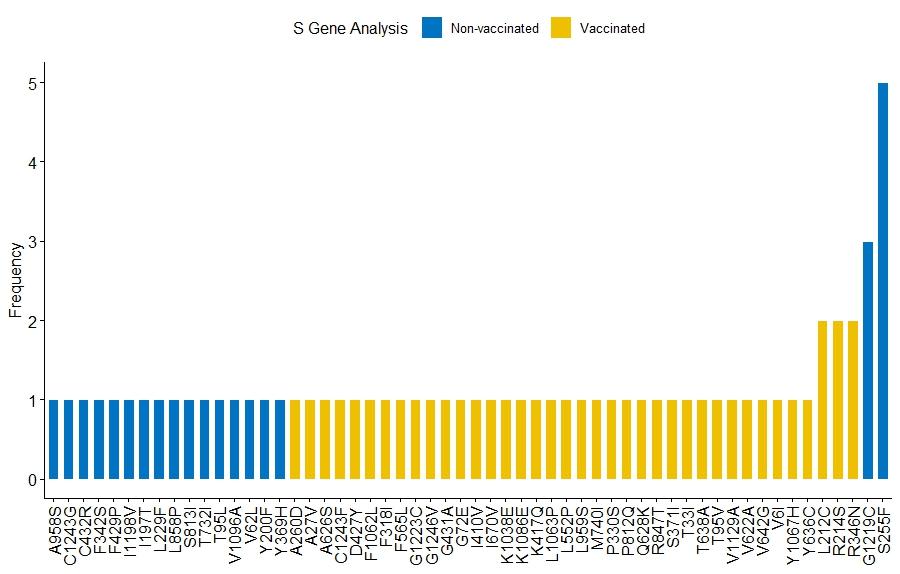

**B.**

**C.**

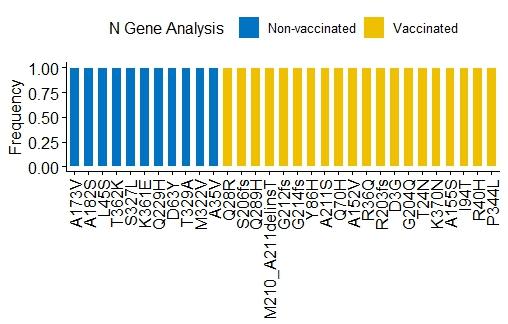

Supplementary Fig. 11: Unique mutations in non-vaccinated and vaccinated patients in Kenya. Mutations in blue represent those found in non-vaccinated patients and those in yellow represent those in vaccinated patients. A. Shows unique mutations in the ORF1 a/b. B. Shows unique mutations in the S genes C. Shows unique mutations in the N gene.
